# Human DNA methylation and the cortisol response to an acute psychological stressor: a systematic review and meta-analysis

**DOI:** 10.1101/2025.10.23.25338702

**Authors:** David Balfour, Zoe Kleinig, Murthy Mittinty, Sarah Cohen-Woods

## Abstract

The hypothalamic-pituitary-adrenal axis (HPA axis) is an important part of the stress response. The HPA axis may adapt to the environment in part through epigenetics, including DNA methylation. This pre-registered (OSF), PRISMA-compliant systematic review and meta-analysis aimed to evaluate the level of evidence for an association between DNA methylation and the cortisol response to an acute psychological stressor, a key marker of HPA axis function, in humans. PsycINFO, MEDLINE, Scopus, and Web of Science were searched on the 1^st^ of September 2025 and risk of bias was evaluated using an original rubric. Thirty-nine studies were included, with mixed results. Meta-analyses revealed support for an association between *NR3C1* methylation and a stronger cortisol response in infants (*r* = .26, *p* = .01), but not other age groups (*r* = -.01, *p* = . 85). There was some tentative evidence for an association between *SLC6A4* methylation and a weaker cortisol response (*r* = -.15, *p* = .056), but the effect was not significant. There was preliminary (non-meta-analytic) support for *LEP*, *NR3C2*, *OXTR*, and *SKA2*. The mixed results may be a product of confounding, small sample sizes, and candidate gene methods. They may also reflect a true but complex relationship, observable only in certain populations (e.g., infants) or in conjunction with other biological or environmental factors (e.g., antidepressants). We propose several directions for research, including a collaborative, pre-registered meta-analysis and a focus on genomic loci that may be more strongly correlated between brain and peripheral tissue. This review received no specific funding.

## Introduction

Stress can have a significant impact on health and emotional wellbeing [1, 2] and has been shown to impose major economic burden through productivity loss and cost of healthcare [3]. Given the high impact of stress, there is a strong need to understand the pathways that underpin its effect. When the individual encounters a stressor, the hypothalamic-pituitary-adrenal axis (HPA axis) works to increase the secretion of cortisol. Cortisol is a powerful hormone that has widespread effects on metabolism, the immune system, cognition, and brain function [4, 5]. Signs of dysregulation within the HPA axis (e.g., high or low cortisol) have been associated with a variety of common diseases, including depression, diabetes, and hypertension [6, 7, 8, 9]. A better understanding of factors that underpin the function of the HPA axis may help to improve our knowledge of these conditions.

The cortisol response to stress is a key marker of HPA axis function, indexing the reactivity of the system [10]. Individuals can show moderate, heightened, or blunted reactivity, which may confer differences in their vulnerability to stress-related pathology [11]. The gold standard method for investigating the cortisol response is the Trier Social Stress Test (TSST) [10]. The TSST is a standard experimental protocol in which participants are asked to complete a 5 minute speaking task followed by a 5 minute mental arithmetic task in front of an unsympathetic panel of researchers. During the procedure, repeated measurements of blood or saliva are obtained to calculate the change in cortisol in response to the psychological stressor. Using the TSST or another acute stressor, researchers can investigate factors that may contribute to or underpin the function of the HPA axis.

Animal studies indicate that the hormonal response to stress may adapt to the environment in part through changes in the epigenome [12, 13]. The epigenome is a collection of mitotically heritable chemical and structural changes to DNA that can affect gene expression without altering the underlying sequence of bases [14]. The most-studied epigenetic mark is DNA methylation, which involves the addition of a methyl group to DNA. According to the epigenetic programming hypothesis, certain environmental influences such as childhood trauma and exposure to maternal stress in utero, may alter DNA methylation, which may in turn influence gene expression and the function of the HPA axis [15, 16, 12], [13]. Altered function of the HPA axis may then have downstream effects on mental and physical health, contributing to observed relationships between stress, markers of HPA axis function, and health [6]. We aimed to evaluate the level of evidence for the proposed relationship between the epigenome and the function of the HPA axis in humans by conducting the first systematic review and meta-analysis of DNA methylation and the cortisol response to an acute psychological stressor.

## Methods

### Eligbility Criteria

This review is based on PRISMA 2020 [17]. A protocol for the review was uploaded to OSF on the 10^th^ of September 2024 [18]. Records were considered eligible if they analysed DNA methylation and the cortisol response to an acute psychological stressor. A stressor was considered acute if it took place over a period of no more than 24 hours. Records were excluded if they were not peer reviewed journal articles or if they were not in English.

### Search Strategy

Search queries included both a term for DNA methylation or epigenetics and a term related to stress, the cortisol response, or the function of the HPA axis (full queries are in the Supplementary Material). Four databases were searched on the 1^st^ of September, 2025 (PsycINFO, MEDLINE, Scopus, and Web of Science) and the results were uploaded to Covidence. Two reviewers (DB and ZK) independently screened the records. Two reviewers (DB and either SCW or ZK) then independently extracted the study characteristics and results (Table 1).

**Table 1.**
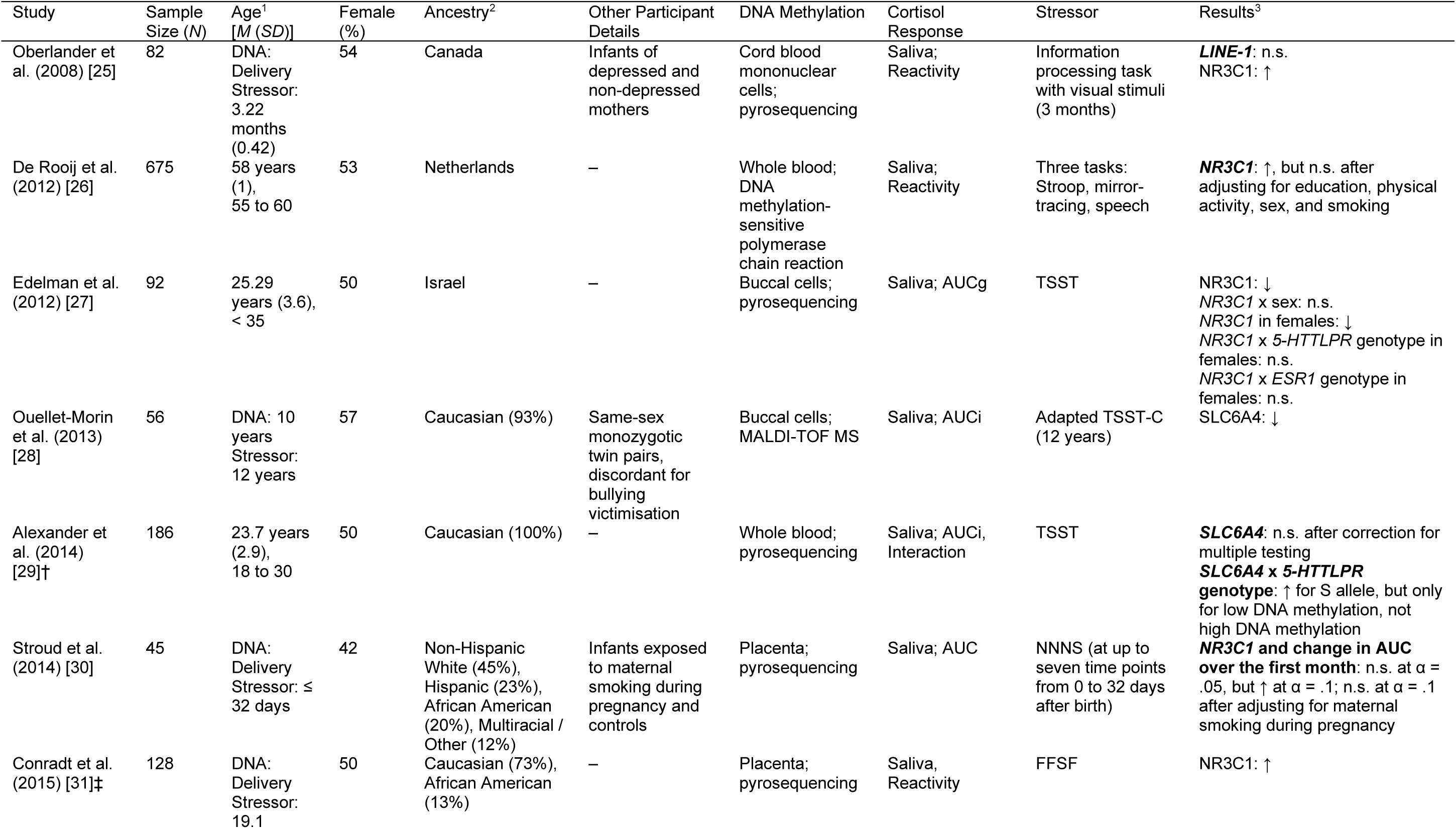
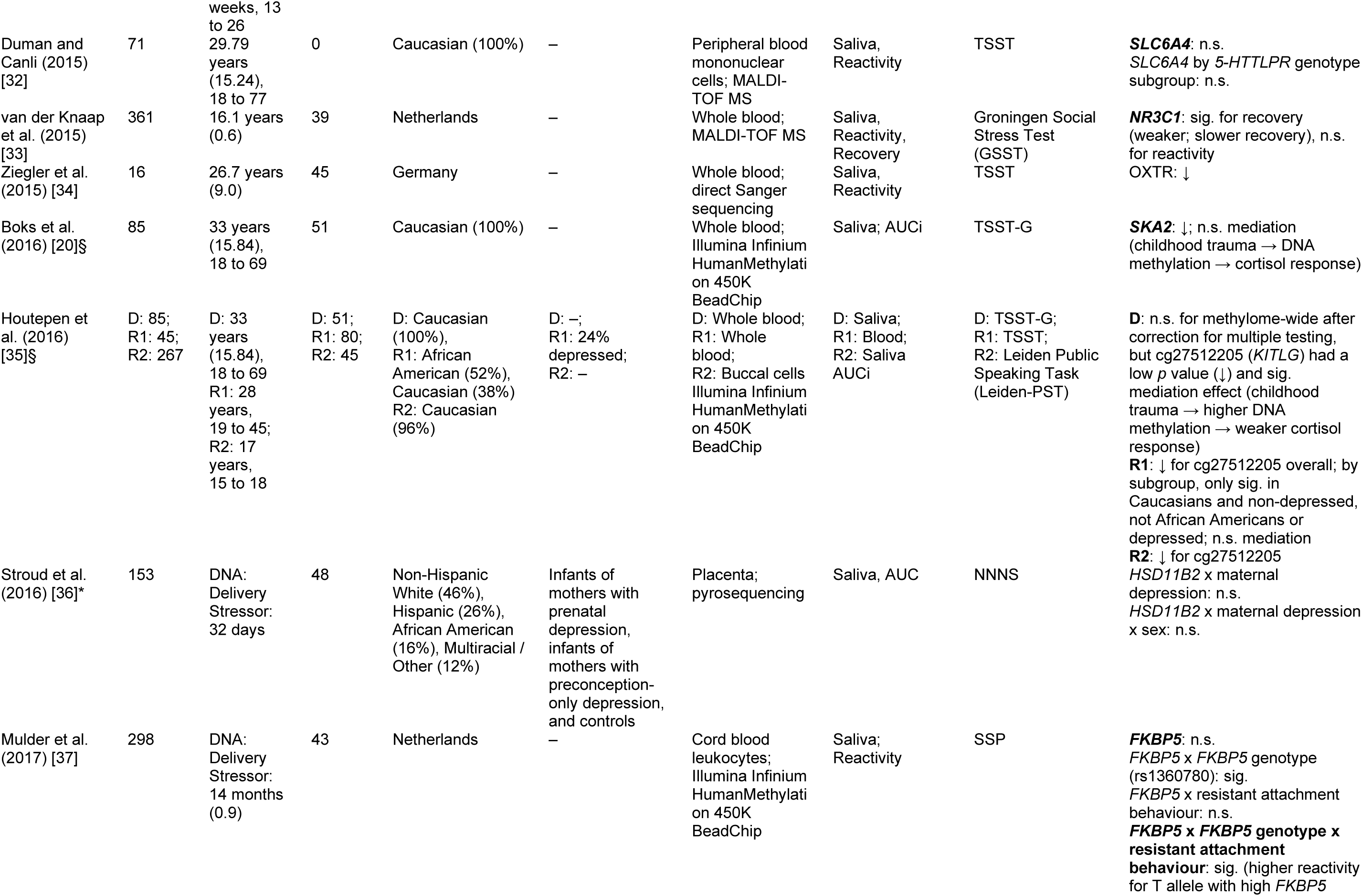
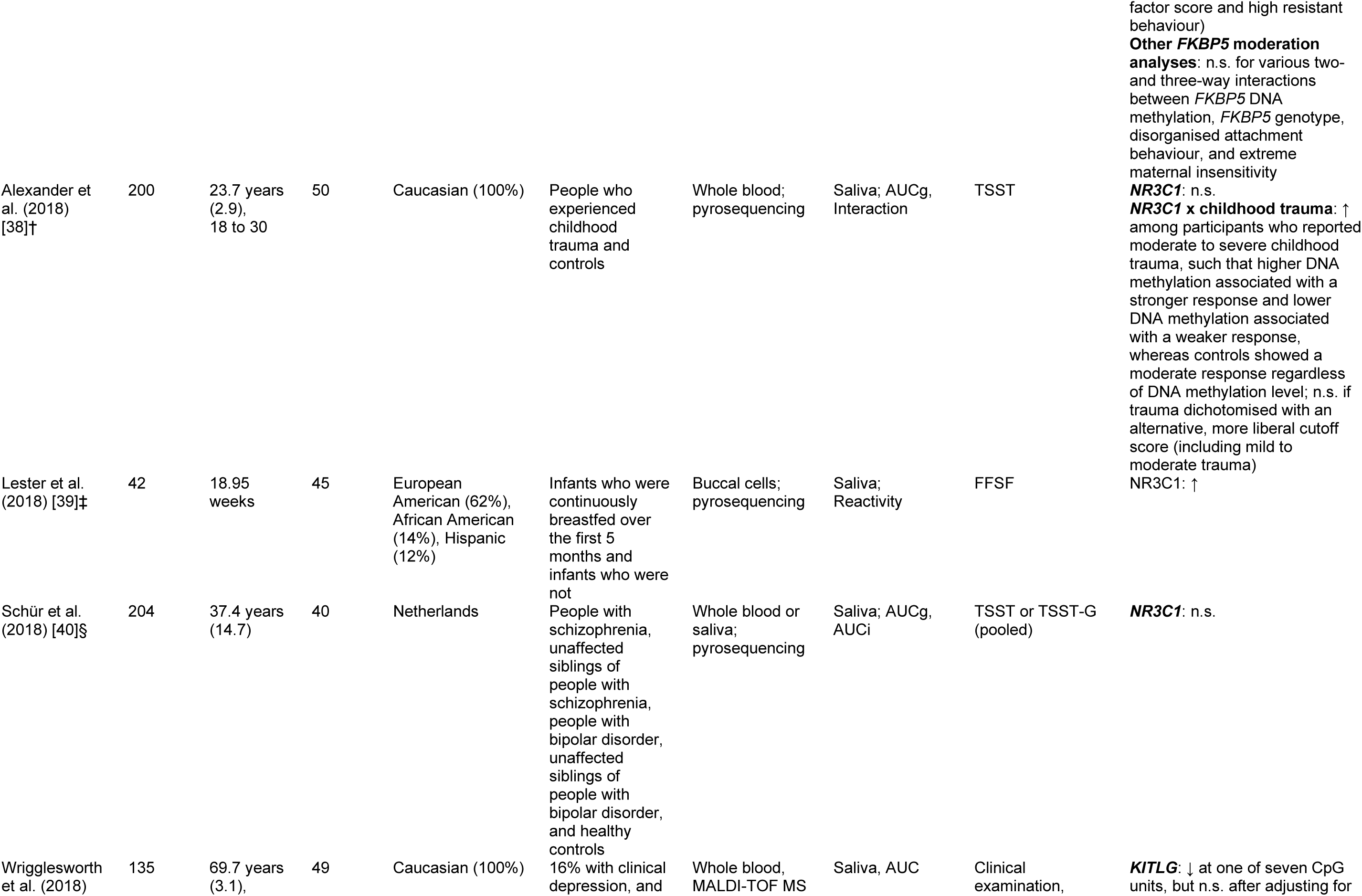
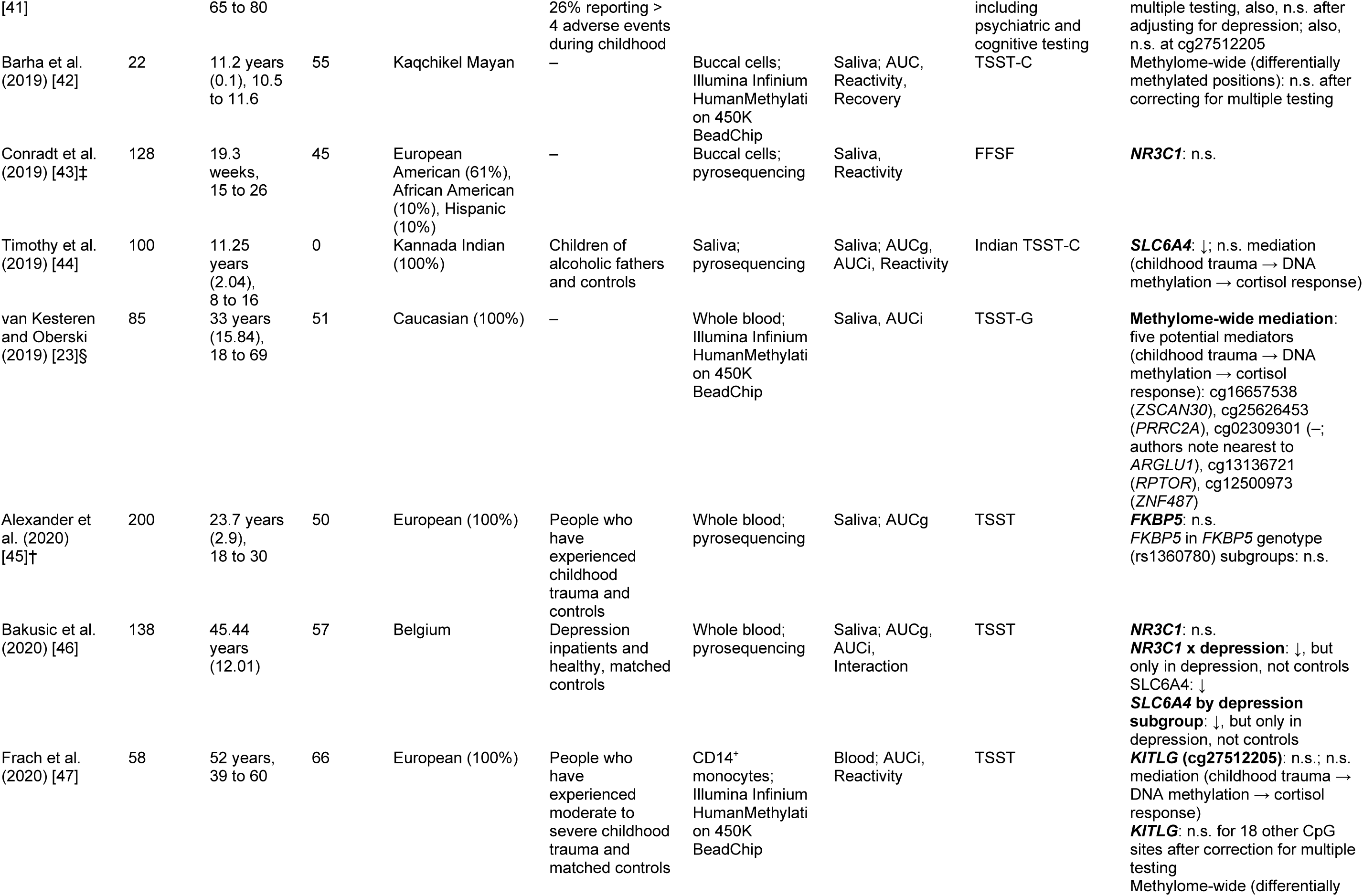
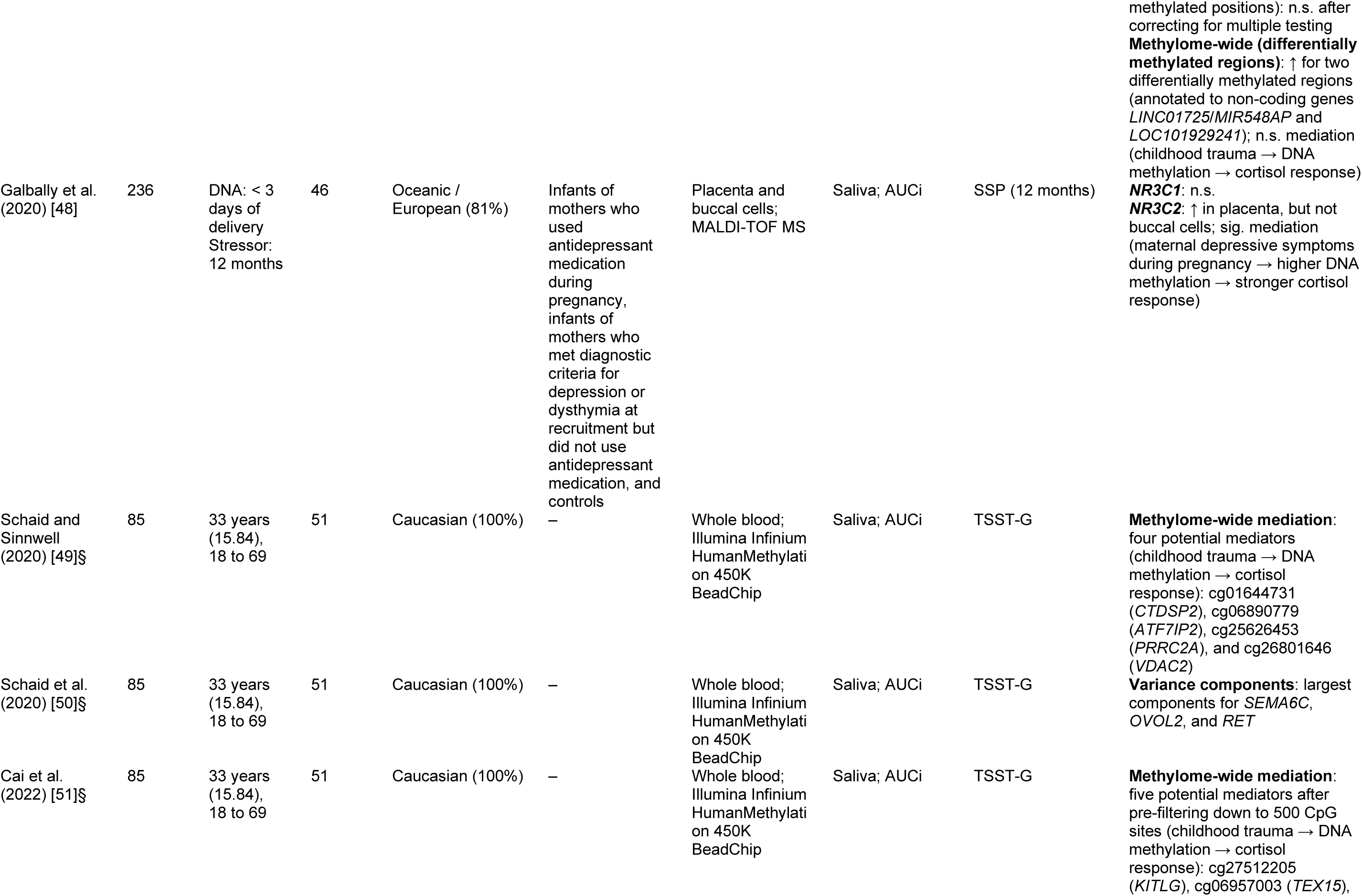
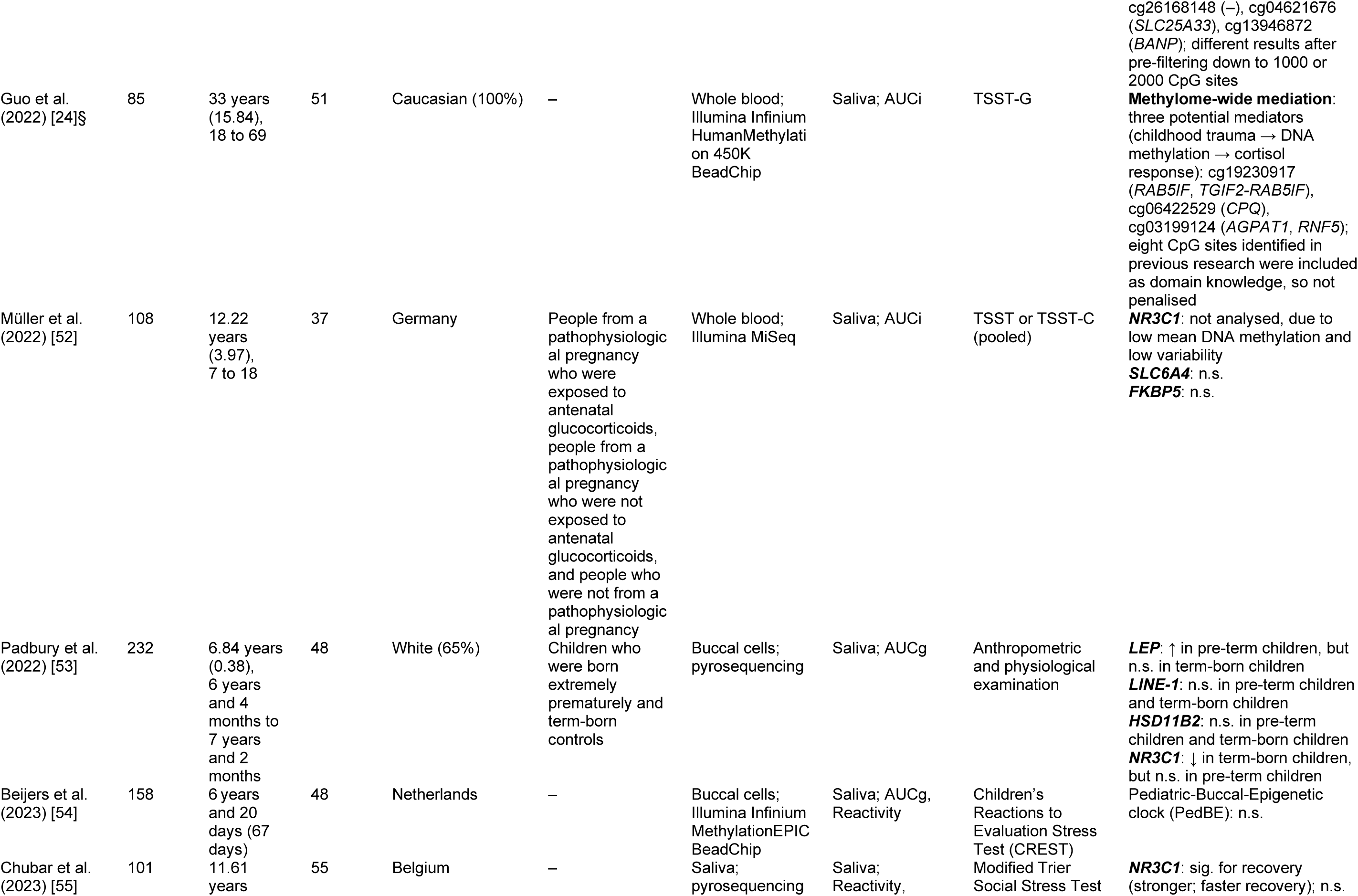
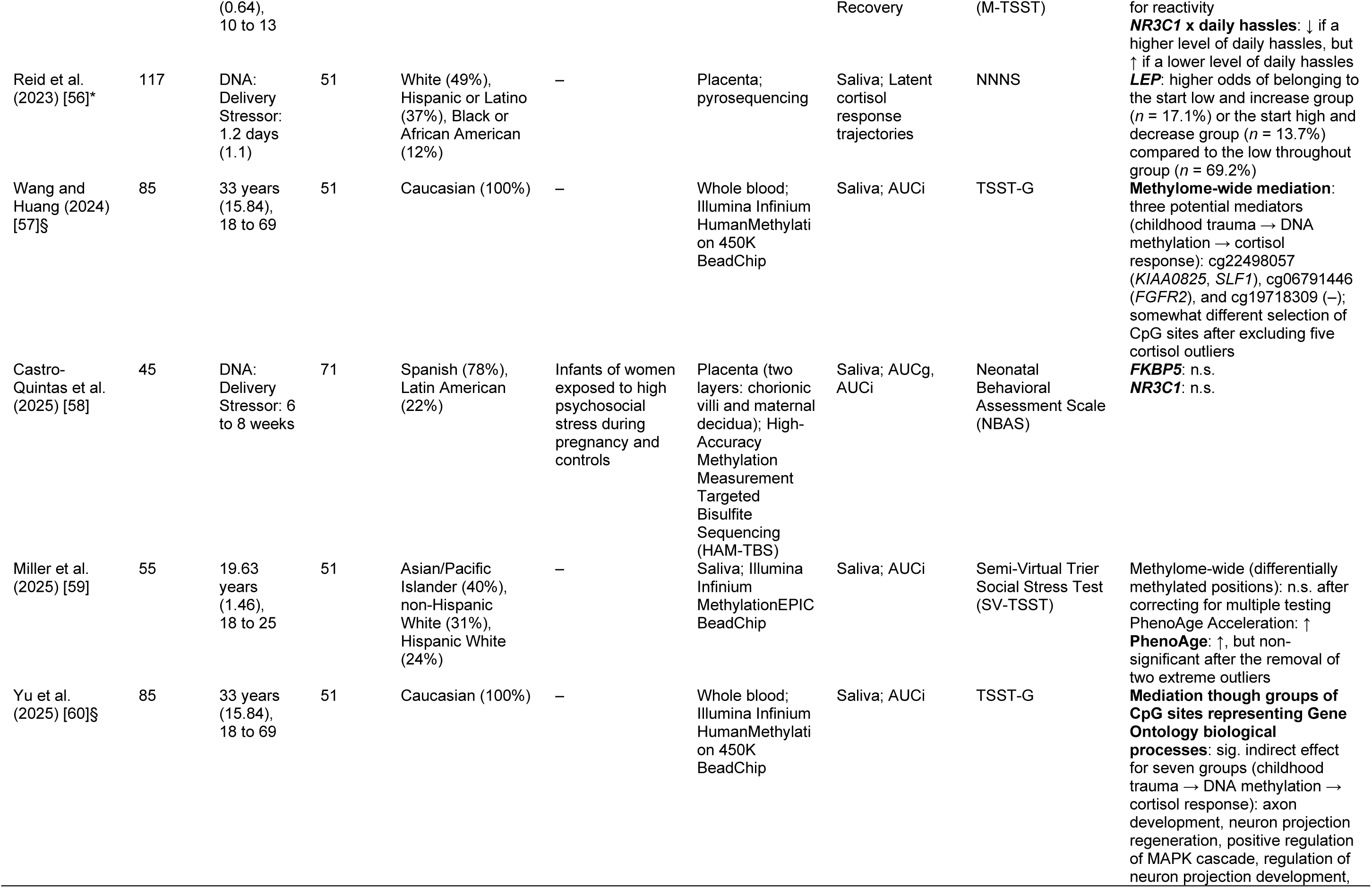
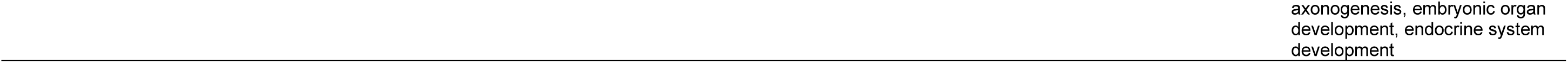
Study Characteristics and Results. ↑ = higher DNA methylation was associated with a stronger cortisol response, ↓ higher DNA methylation was associated with a weaker cortisol response, – = not applicable, D = discovery cohort, R1 = replication cohort 1, R2 = replication cohort 2, FFSF = Face-to-Face Still Face, interaction = interaction between time and DNA methylation, MALDI-TOF MS = matrix-assisted laser desorption/ionisation time-of-flight mass spectrometry, NNNS = Neonatal Intensive Care Unit Network Neurobehavioral Scales, SSP = Strange Situation Procedure, TSST = Trier Social Stress Test, TSST-C = Trier Social Stress Test for Children, TSST-G = Trier Social Stress Test for Groups. Symbols in the “Study” column (*, †, ‡, §) indicate samples that included the same participants or may have done so. Some participant characteristics are approximate, based on a larger sample than the one used in the relevant analyses. ^1^Age at the time of the stressor unless otherwise specified ^2^Groups comprising ≥ 10% of the sample; location of recruitment if ancestry not clear ^3^CpG sites in this column are accompanied by annotated protein-coding genes in brackets; “–” indicates that the site is not annotated to a protein-coding gene

### Synthesis Methods

#### Synthesis

Meta-analyses were conducted if there were three or more estimates for the same gene and measure of the cortisol response: reactivity, recovery, area under the curve with respect to the ground (AUCg), or area under the curve with respect to the increase (AUCi). Here, “reactivity” refers to any measure of the magnitude and direction of the *initial* change in cortisol on exposure to the stressor (usually the baseline value subtracted from the peak) and “recovery” refers to any measure of the magnitude and direction of the change in cortisol after the peak. By contrast, “cortisol response” refers to *any* measure of change in relation to the stressor.

#### Meta-Analyses

Meta-analyses were performed with the package *metafor* in R version 4.5.1. Random effects models were used to account for between-study heterogeneity. Additional levels were added to account for the lack of independence between effects for different genomic loci in the same participants (e.g., different amplicons in the same gene) and partly overlapping cohorts between some studies. This multilevel approach involves minimal assumptions and has been shown to perform well in simulation research [19]. Forest plots depict Fisher’s *r*-to-*z* transformed correlation coefficient, with Pearson’s *r* in the text. Heterogeneity was quantified using *I^2^*and sources of heterogeneity were explored with subgroup analyses and meta-regression, as described in the results. For the AUCi meta-analyses, if prior publications had not reported specifically on the gene of interest in the publicly available data set GSE77445 [20], we computed original estimates using those data and included them to increase statistical power and provide additional information about the relationship of interest (referred to as “Balfour et al., 2025” in the forest plots). Full quantitative methods are available in the Supplementary Material.

### Assessment of Risk of Bias

We generated a new risk of bias rubric to capture the most relevant areas of potential concern (Table S1). The rubric was based on the Q-Genie [21], but developed specifically for epigenetic rather than genetic research. Two reviewers (DB and SCW) performed the initial risk of bias assessment independently, assigning one of three levels for each domain in each study. The reviewers then resolved any discrepancies that exceeded one level through discussion. The final risk of bias judgements were defined as the mean score between the reviewers, rounded up. Following the pre-registered protocol [18], funnel plots were not used to assess publication bias, as there were no meta-analyses that included effects from more than 10 studies. As noted by Sterne et al. [22], funnel plots that include fewer than 10 effects are not likely to be able to differentiate between real and chance asymmetry.

## Results

### Study Selection

The study screening and selection processes are reported in Figure 1.

**Figure 1.**
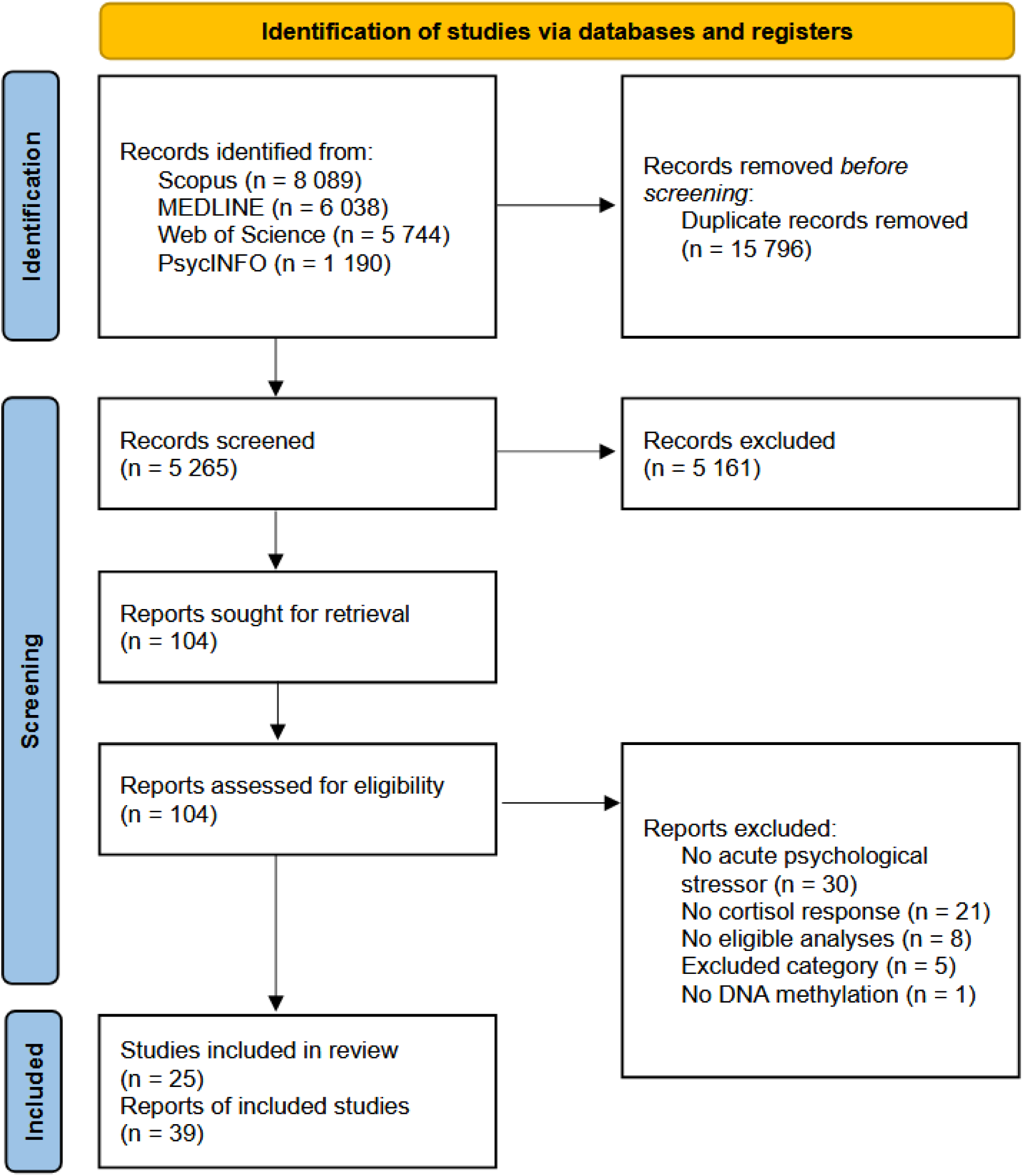
PRISMA Flow Diagram. Following PRISMA, studies are distinct investigations that include a defined group of participants and one or more interventions or outcomes. By contrast, reports are documents that provide information about a study – in some cases, one study may have multiple reports [17].

Thirty-nine reports of 25 distinct studies were included. One report [23] was identified through a reference [24], not the systematic search.

### Study Characteristics

Twenty-eight reports included at least one candidate gene analysis, and four included a methylome-wide analysis, to identify the main effect of differentially methylated positions. Study characteristics and results are reported in Table 1.

### Results of Syntheses

#### NR3C1

*NR3C1* was the most popular target gene, analysed using a candidate gene method in 15 studies. The findings were mixed, with four studies reporting evidence for an association with a stronger cortisol response [31, 26, 39, 25], one with a weaker response [27], one with stronger (faster) recovery [55], and one with weaker (slower) recovery [33]. Six studies reported no evidence for a main effect of *NR3C1* methylation [38, 46, 43, 48, 40, 30] (at α = .05). Interaction effects were reported between methylation at the gene and childhood trauma [38], recent daily hassles [55], and depression [46]. Our meta-analyses for reactivity (Figure 2, *k* = 12, *N* = 1 389), recovery (Figure S1, *k* = 6, *N* = 462), AUCg (Figure S2, *k* = 7, *N* = 911), and AUCi (Figure S3, *k* = 4, *N* = 472) revealed no evidence for an overall effect of *NR3C1* methylation (absolute *r* ≤ .09 and 95% confidence intervals included zero).

**Figure 2.**
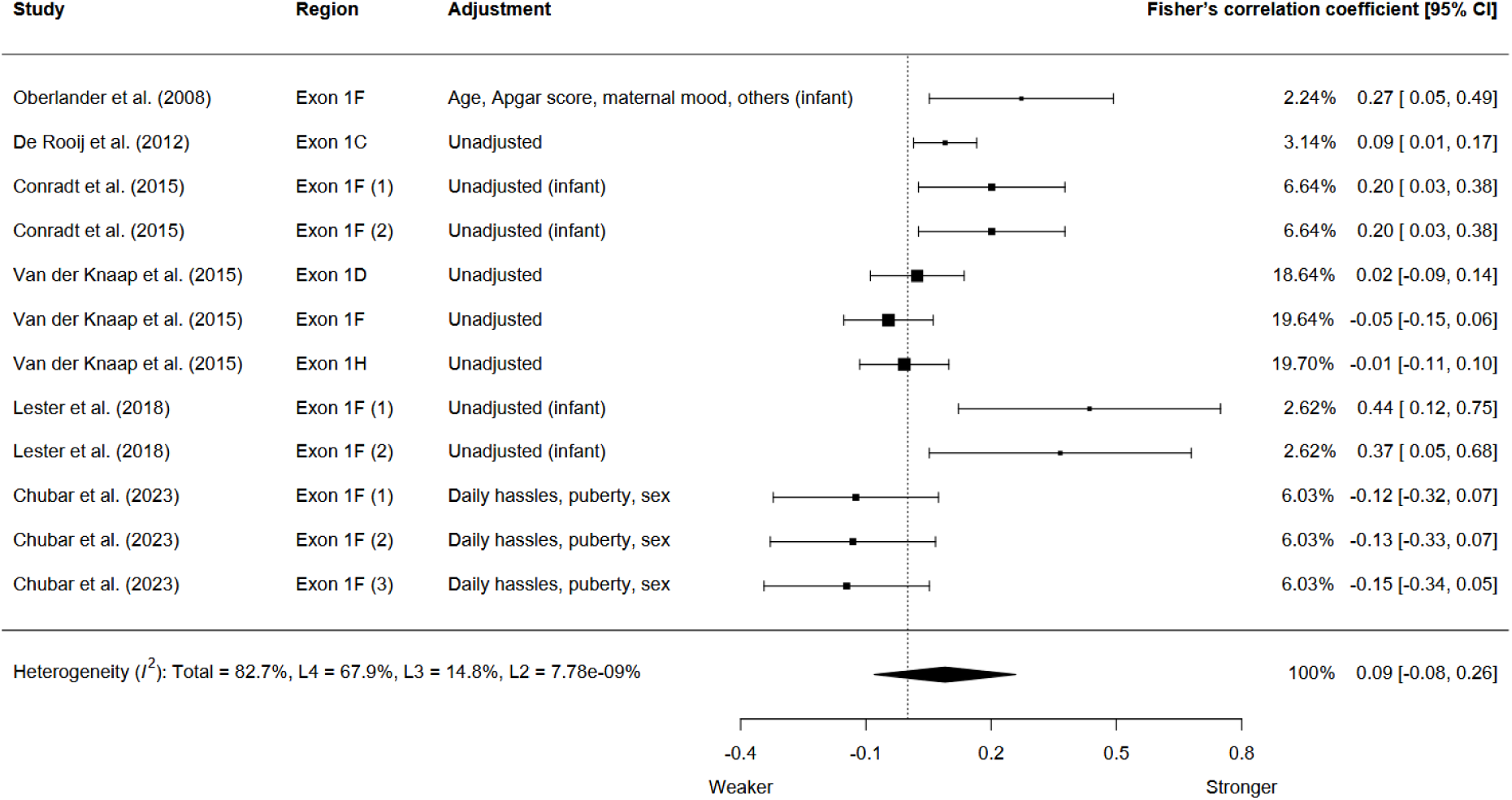
Association Between *NR3C1* Methylation and the Cortisol Response (Reactivity) Pooled estimate: *r* = .09 (95% CI = -.08, .25), *p* = .27. Weaker = weaker cortisol response, Stronger = stronger cortisol response. Region column indicates the specific area within *NR3C1*, if the authors reported on more than one for the same participant group. Adjustment column indicates which variables were adjusted for, with the word “infant” in brackets to indicate an infant-only sample. Additional levels included in the meta-analysis to account for the lack of independence between estimates for different genomic loci within studies and partially overlapping cohorts between studies.

#### SLC6A4

*SLC6A4* was the second most popular target gene, analysed using a candidate gene method in six studies. The results were mixed, with three studies reporting a relationship between *SLC6A4* methylation and a weaker cortisol response [46, 28, 44] and three reporting no evidence for a main effect of methylation at the gene [29, 32, 52]. One study reported evidence for an interaction between *SLC6A4* methylation and *5-HTTLPR* genotype [29], but a later study was unable to replicate the effect [32]. One study reported no evidence for an indirect effect of childhood trauma on the cortisol response through *SLC6A4* methylation [44]. Our meta-analyses revealed tentative, preliminary support for an association between *SLC6A4* methylation and a weaker cortisol response, both as AUCg (Figure S4, *k* = 2, *N* = 238, *r* = -.21 [95% CI = -.27, -.16], *p* = .01) and AUCi (Figure 3, *k* = 6, *N* = 673, *r* = -.15 [95% CI = -.3, .01], *p* = .056). However, the estimate for AUCg was only based on two studies and the 95% confidence interval for AUCi included zero.

**Figure 3.**
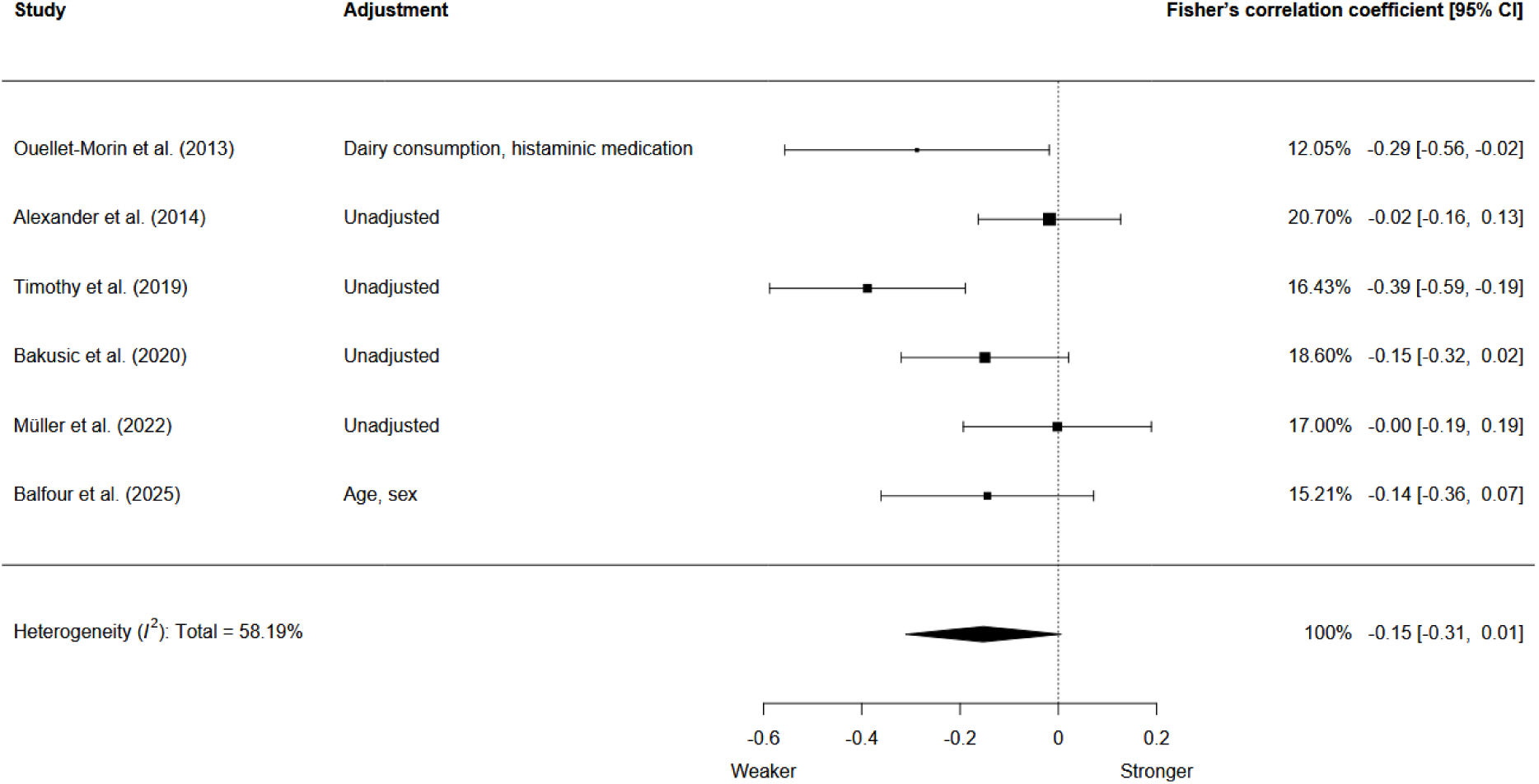
Association Between *SLC6A4* Methylation and the Cortisol Response (AUCi) Weaker = weaker cortisol response, Stronger = stronger cortisol response. Adjustment column indicates which variables were adjusted for.

#### Other Genetic Loci

Four studies targeted *FKBP5*, and none reported evidence for a main effect of DNA methylation on AUCg, AUCi, or reactivity [45, 58, 37, 52]. Meta-analysis also revealed no evidence for an effect of *FKBP5* DNA methylation (absolute *r* ≤ .04 and 95% confidence intervals included zero; AUCi: Figure S5, *k* = 5, *N* = 185; AUCg: Figure S6, *k* = 4, *N* = 245).

Some genomic loci were targeted by two studies each. For *HSD11B2* and LINE-1 (a marker of global DNA methylation), no studies reported evidence for an association (*N* = 232 [53], *N* = 153 [36], *N* = 82 [25], *N* = 232 [53]). The two studies that investigated *LEP* reported evidence for an association between DNA methylation at the gene, and a more responsive HPA axis [53, 56]. Two studies investigated age-related DNA methylation profile scores. Of the two, one reported no evidence for an effect of the paediatric buccal cell-based model PedBE (AUCg, *N* = 158) [54] and the other reported an association between the blood-based model PhenoAge and a stronger cortisol response (AUCi, *N* = 55) [59].

Some genomic loci were targeted by one study each. An association was reported between DNA methylation of *OXTR* and a weaker cortisol response (reactivity) (*N* = 16) [34]. An association between DNA methylation of *SKA2* and a weaker cortisol response (AUCi) was observed, although DNA methylation did not act as a mediator between childhood trauma and cortisol response (*N* = 85) [20]. Finally DNA methylation of *NR3C2* was associated with a stronger cortisol response in infants in placenta but not in infant buccal cells (AUCi, *N* = 236) [48], and placental *NR3C2* methylation mediated between maternal depressive symptoms in early pregnancy and infant cortisol response [48].

#### Methylome-Wide Analyses

There were four independent methylome-wide studies. In the first and largest (*N* = 85), Houtepen et al. [35] reported no significant effects after correction for multiple testing, although one CpG site in *KITLG* (cg27512205) was nominally associated with a weaker cortisol response (AUCi), and this effect replicated in two other cohorts (*N* = 45 and *N* = 267). The same CpG site also strongly mediated the relationship between childhood trauma and cortisol response, although this effect did not replicate. Two independent studies targeted the CpG site in *KITLG* previously identified by Houtepen et al. [35] but failed to replicate the effect (*N* = 135 [41], *N* = 58 [47]); one of the two reported a relationship between DNA methylation at a nearby CpG unit in *KITLG* (one of seven examined) and a weaker cortisol response (AUCi), but the effect became non-significant after adjusting for depression [41]. Our meta-analysis for the CpG site in *KITLG* identified by Houtepen et al. [35] revealed a substantial point estimate given the context, but the 95% confidence interval included zero (AUCi; Figure S7, *k* = 4, *N* = 455, *r* = -.28 [95% CI = -.55, .04], *p* =.07). Three other methylome-wide studies also reported no differentially methylated positions after correction for multiple testing (*N* = 58 [47], *N* = 55 [59], *N* = 22 [42]).

### Heterogeneity

#### Evidence of Heterogeneity

There was evidence for relative heterogeneity (*I*^2^), with approximately 58 to 83% of the variance in the largest meta-analyses for *NR3C1* and *SLC6A4* being attributable to possible differences in the true effect between cohorts (Figure 2 and Figure 3).

#### Participant Age

Because infants show age-related differences in the cortisol response [61], we performed subgroup analyses by infant status. The point estimate for *NR3C1* methylation and reactivity (Figure 2) more than doubled in strength and became significant after restricting the analysis to infants (Figure 4A, *k* = 5, *N* = 252, *r* changed from .09 to .26 [95% CI = .1, .4] and *p* changed from .27 to .01), but it moved close to zero in non-infants (Figure 4B, *k* = 7, *N* = 1 137, *r* = -.01 [95% CI = -.16, .14], *p* = .85). Meta-regression revealed evidence for an effect of infant status, *t*(10) = 3.08, *p* = .01. Sufficient data were not available to perform age-specific analyses for the other genes.

**Figure 4.**
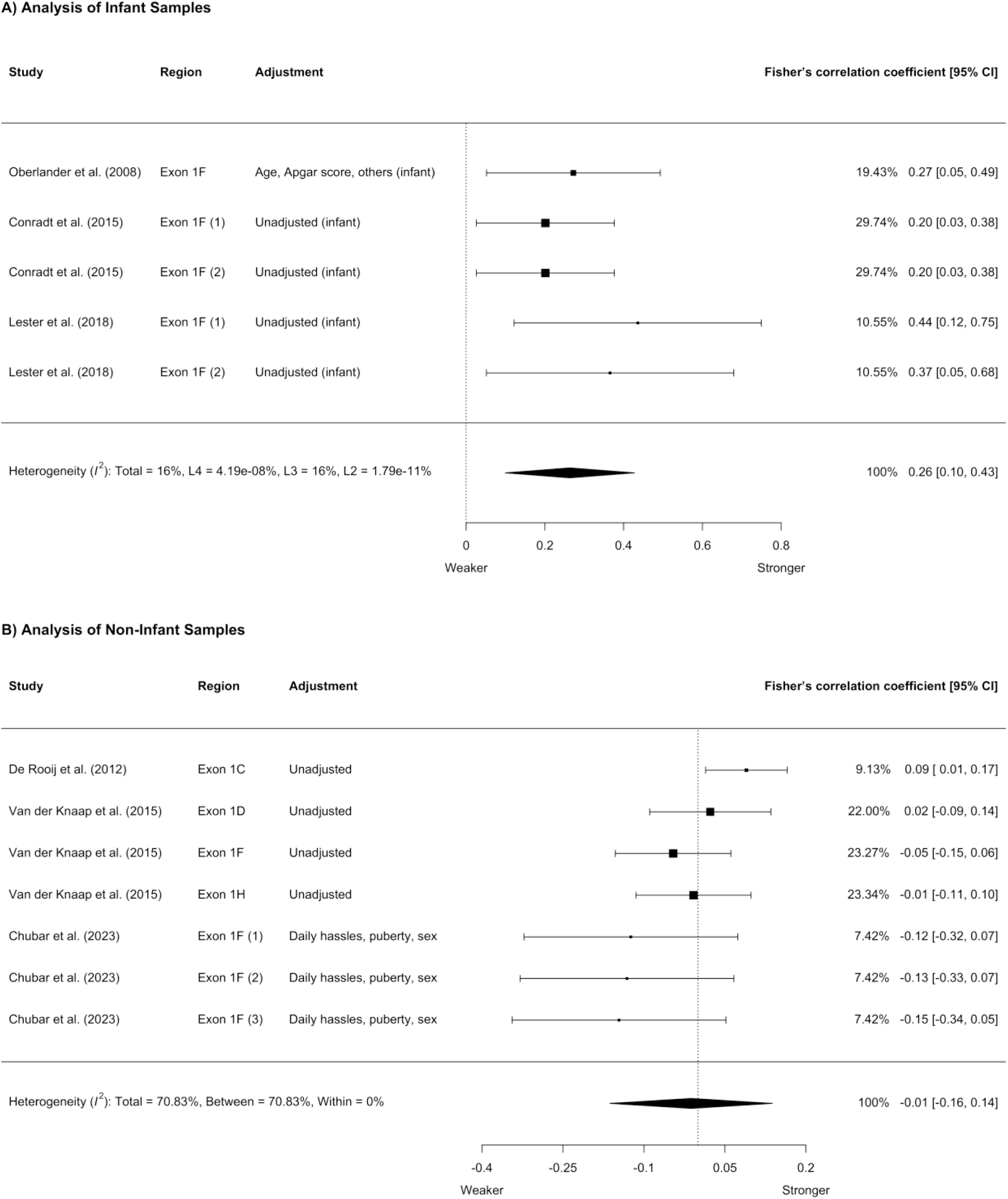
Association Between *NR3C1* Methylation and the Cortisol Response (Reactivity) in Infants and Non-Infants. Region column indicates the specific area within *NR3C1*, if the authors reported on more than one for the same participant group. Adjustment column indicates which variables were adjusted for, with the word “infant” in brackets to indicate an infant-only sample. Additional levels included in the infant meta-analysis to account for the lack of independence between estimates for different genomic loci within studies and partially overlapping cohorts between studies.

#### Selection of Genetic Loci

For *NR3C1*, studies investigated different splice variants of exon 1 (1_C_, 1_D_, 1_F_, and 1_H_). Exon 1_F_ was the most popular in part because there is evidence for an effect of its rodent homologue, exon 1_7_, on the glucocorticoid response to stress. To account for the possibility of an effect specific to exon 1_F_, we performed meta-analyses that excluded the other exons. The pooled estimates did not change substantially (reactivity: Figure S13; recovery: Figure S14; AUCg: Figure S15). We did not have sufficient data to perform region-specific meta-analyses for the other genes. See the Supplementary Material for additional comments on heterogeneity.

### Risk of Bias

Risk of bias judgements are reported in Table S2. These identified some concerns about selection of participants, with potential for confounding by distinct subgroups (e.g., participants with depression and controls). Most candidate gene studies provided a clear rationale [62], [63], however the methylome-wide studies were not as well justified given their small sample sizes. Power analyses for the largest methylome-wide study (*N* = 85) [35] indicate that power was not likely to have been above .0205 (Table S3; details in the Supplementary Material).

There were significant concerns about the selection of statistical methods. Often, decisions that may have affected the results lacked a strong rationale. In some cases, researchers did not adjust for multiple testing despite a high number of tests, relating multiple, partly independent DNA methylation variables (e.g., CpG sites or amplicons) to multiple, partly independent measures of the cortisol response (e.g., AUCg and reactivity). Other possible issues included – without a clear rationale – use of an unconventionally liberal α level, extensive interaction and subgroup analyses, and dichotomisation of continuous variables using a median split. While the median split is common, it can negatively impact power and increase rate of both type one and type two error [64]. There were substantial concerns regarding the selection of confounders, with some studies failing to adjust for variables that are known to be associated with both DNA methylation and the cortisol response, including age, ancestry, and sex [65, 66, 67]. Studies also often failed to adjust for cell type proportions, which are a common confounding variable in DNA methylation research [68]. In some studies, there was no clear rationale for the covariates that were included.

There were also some concerns about the selection of reported results. While this cannot be assessed with certainty, we interpreted absence of analyses that were most likely performed (based on the available data and the analyses that *were* reported) as evidence of possible selective reporting. Also, studies often lacked sensitivity analyses for potentially important statistical decisions, including choice of covariates and exclusion of a substantial number of participants. In some cases, authors failed to report an effect for all available genetic loci without a clear rationale. The conclusions were usually appropriate and measured, although some causal claims may have been too strong.

## Discussion

The evidence for a relationship between DNA methylation and the cortisol response to an acute psychological stressor is mixed. In methylome-wide research, the most notable finding was for a single CpG site in *KITLG* (cg27512205) [35], which – consistent with epigenetic programming – mediated between childhood trauma and the cortisol response. Follow-up analyses failed to replicate this effect, however [47, 41], and our meta-analysis for the identified CpG site did not show a significant effect. The point estimate for the pooled effect was nevertheless substantial, and further analyses in a larger cohort are required. In candidate gene research, the most popular target was *NR3C1*, in part due to evidence for *Nr3c1* methylation as a potential mediator between differences in maternal care and the glucocorticoid response to stress in rodents [13]. Findings for *NR3C1* have varied substantially and our meta-analyses revealed no support for an effect overall, suggesting the findings from animal research may not generalise neatly to humans.

The mixed results for *NR3C1* may be the product of a true but complex relationship, observable only in conjunction with other biological or environmental factors. Consistent with this possibility, interactions were reported between *NR3C1* methylation and depression [46], childhood trauma [38], and recent daily hassles [55], although these have not yet been replicated. Age may also be a relevant factor, with meta-regression revealing evidence for an effect of *NR3C1* methylation on the cortisol response in infants (Figure 4A) but not in other age groups (Figure 4B). Consistent with the effect observed in rodents, higher levels of DNA methylation were associated with a stronger cortisol response [25, 69]. One possible interpretation is that infancy may be a critical period for the adaptation of the HPA axis to the environment through *NR3C1* methylation, with accumulated environmental exposures (e.g., those associated with different lifestyles and social experiences) becoming more powerful determinants or correlates of the cortisol response over time. Despite the plausibility of the observed effect, it is important to note that our infant meta-analysis cannot provide more than weak, early support, as it only included three studies. Also, human research has been limited to peripheral tissue, whereas the effect observed in rodents was for hippocampal DNA methylation, which is more directly relevant because the hippocampus facilitates negative feedback within the HPA axis. Future studies may be able to overcome this limitation to some extent by focusing on CpG sites that may be more strongly correlated between brain and peripheral tissue [70, 71, 72].

Our meta-analyses revealed some evidence for a relationship between *SLC6A4* methylation and a weaker cortisol response. However, contrary to the notion of epigenetic programming, the only study that investigated *SLC6A4* methylation as a potential mediator between an environmental exposure (early adversity) and the cortisol response reported no evidence for an effect (*N* = 100) [44]. Also, our largest meta-analysis for *SLC6A4* cannot provide more than weak, nominal support, as the 95% confidence interval included zero (*p* = .056, Figure 3) and the largest contributing effect was at high risk of confounding due to distinct subgroups representing the children of alcoholic fathers and controls [44]. More specifically, the children of alcoholic fathers had both higher DNA methylation and a weaker cortisol response than controls, which could be expected to produce the observed relationship in the total sample. Future studies that include distinct subgroups should consider adjusting for group status to mitigate any potential confounding.

Some findings were consistent with possible confounding due to depression or serotonin reuptake inhibitors, which are associated with DNA methylation [73] and the function of the HPA axis [74]. In one study, the effect of *SLC6A4* methylation only held in participants with depression, not in controls [46]. In the largest methylome-wide study to date, the effect reported for *KITLG* only held in participants *without* depression, but not in participants with the disorder [35]. A later study reported an effect for a particular CpG unit in *KITLG*, but it became non-significant after adjusting for depression [41]. Future studies should adjust for depression or antidepressant use to help elucidate the possible role of these variables in the relationship of interest.

When interpreting these results, it is important to consider the effect of cortisol on DNA methylation. When the cortisol-receptor complex binds to a glucocorticoid response element, it can cause rapid local demethylation [75]. It is possible, therefore, that some of the observed effects may be due in part or entirely to an existing difference in the function of the HPA axis rather than epigenetic programming (i.e., an effect of DNA methylation). Consistent with this possibility, several studies have reported evidence for an effect of acute psychological stress on DNA methylation [76, 59, 77].

There are various subtleties of the cortisol response to consider. A weaker response, for example, may be interpreted as evidence for a less sensitive HPA axis, but it could also indicate a *more* sensitive HPA axis if it indicates an anticipatory response to the stressor, resulting in a higher level of cortisol at baseline and a less positive change after exposure to the stressor [78, 79, 37]. AUCg and AUCi are conceptually and statistically distinct, with the former representing the total concentration of cortisol throughout the stressor (i.e., relative to zero) and the latter representing the change in response to the stressor relative to the baseline level [80, 81]. Recovery variables may capture a distinct aspect of the function of the HPA axis (i.e., the suppression of the initial cortisol response), although few studies included them [82]. We encourage researchers to perform sensitivity analyses with different cortisol response variables and adjustment for the baseline value to investigate the robustness of their results and explore any potentially meaningful differences in the effect. In general, this will not require the collection of additional data.

Several limitations must be kept in mind. First, most studies in this review used candidate gene analyses, which tend to produce less reliable findings [62, 63]. Unfortunately, methylome-wide analyses require a stringent correction for multiple testing, limiting statistical power. As noted in the section on risk of bias, the reviewed methylome-wide analyses were severely underpowered. Also, due to the limited availability of data, it was only possible to perform meta-analyses for four genes (*NR3C1*, *SLC6A4*, *FKBP5*, and *KITLG*). Most of these meta-analyses were small, in part because different studies that investigated the same gene used different cortisol response variables. Also, it is important to note that the limited availability of data may have introduced bias into our meta-analyses, as significant findings were sometimes more completely reported and therefore more available for inclusion than non-significant ones. Issues with limited statistical power and incomplete reporting may be addressed through a pre-registered collaborative meta-analysis, similar to the one conducted by the Psychiatric Genomics Consortium for post-traumatic stress disorder [83, 84]. The collaborators could follow a specific analytical protocol to ensure conceptually comparable effects between the cohorts (e.g., with the same cortisol response variable and covariates).

It is not yet possible to draw any strong conclusions regarding the relationship between DNA methylation and the cortisol response to an acute psychological stressor in humans. The mixed results may be the consequence of a true but complex effect, observable only in certain populations (e.g., infants) or in conjunction with other biological or environmental factors (e.g., antidepressants). Alternatively, they may be the result of sampling error, confounding, or a reliance on candidate gene methods. The observed relationships could also be the result, in part, of an existing difference in the function of the HPA axis. We propose several directions for research, including a collaborative meta-analysis with a specific, pre-registered protocol, sensitivity analyses with different strategies for investigating the cortisol response (e.g., adjustment for the baseline value), and a focus on genomic loci that may be more strongly correlated between brain and peripheral tissue.

## Data Availability

The datasets generated and analysed during the current study are available from the corresponding authors on reasonable request.

## Acknowledgements

We would like to thank Shannon Brown, the Senior Librarian in the Research Engagement Team at Flinders University, for her help with the formal search strategy. We would also like to thank the various authors who helped us complete this review by providing more information about their research on request. Thank you, also, to the statistical consultant Dr Pawel Skuza at Flinders University for his feedback on the analyses in the draft manuscript.

## Author Contributions

**David Balfour**: Conceptualisation, Methodology, Validation, Formal Analysis, Investigation, Writing - Original Draft, Writing - Review & Editing, Visualisation, Project Administration. **Zoe Kleinig**: Validation, Investigation, Writing - Review & Editing. **Murthy Mittinty**: Writing - Review & Editing, Supervision. **Sarah Cohen-Woods**: Conceptualisation, Methodology, Validation, Investigation, Writing - Review & Editing, Supervision, Project Administration

