## Supplementary Material for "Human DNA methylation and the cortisol response to an acute psychological stressor: a systematic review and meta-analysis"

David Balfour, Bachelor of Psychology (Honours)[a]; Zoe Kleinig, Bachelor of Psychology (Honours)[a]; Murthy Mittinty, PhD (Mathematical Statistics)[b]; Sarah Cohen-Woods, PhD (Social Genetic and Developmental Psychiatry)[a, c, d]

[a]College of Education, Psychology and Social Work, Flinders University, Bedford Park, South Australia, Australia

[b]College of Medicine and Public Health, Flinders University, Bedford Park, South Australia, Australia

[c]Flinders Centre for Innovation in Cancer, Bedford Park, South Australia, Australia

[d]Flinders University Institute for Mental Health and Wellbeing, Flinders University, Bedford Park, South Australia, Australia

**Registration and Protocol**

The review protocol was uploaded to OSF on the 10th of September 2024 [1]. An earlier version of the protocol was uploaded on the 15th of August 2024, but it was subsequently revised. The review originally had a narrower focus, on psychosocial rather than psychological stressors. Searches were performed, and – during study screening – we decided to broaden the scope to include all psychological stressors. This was done in part because it was not clear if some of the stressors were psychosocial, although they were clearly psychological. For example, the infant Face-to-Face Still Face paradigm can induce psychological stress through a change in the social environment. However, it was not clear if or how the concept of social-evaluative threat may apply to infants. The broader scope unambiguously classified the Face-to-Face Still Face paradigm as eligible for inclusion, because it is clearly psychological rather than physical or physiological. The broader scope also allowed for the inclusion of more evidence, but the theoretical implications are very similar. We adhered to the updated review protocol, although we added information to the meta-analyses by computing additional effects where possible using the publicly available data set GSE77445 [2]. Also, the search was broadened further, as described below.

**Search Queries**

The final search queries were broader than those in the updated protocol. Specifically, the following phrase was added to each query, to ensure we captured articles that used a range of additional potentially relevant stress protocols: “OR nnnt OR nnns OR ffsf OR "neurobehavioural scale*" OR pasat OR "serial addition task" OR "still-face" OR "still face" OR "evaluation stress test" OR crest OR "neurobehavioural exam*"”. The final search queries are presented below.

***APA PsycINFO***

(((methylation OR dnam OR epigenetic* OR epigenom*).ab,id,ti,tm,hw.) AND ((cortisol adj2 (reactivity OR response)) OR (("hpa axis" OR hpaa OR "hypothalamic pituitary adrenal") adj5 (function OR response OR reactivity OR regulation)) OR (stress* adj2 (acute OR test OR task OR response OR reactivity OR regulation OR psychological OR psychosocial OR social OR mental OR cortisol)) OR tsst OR trier OR groningen OR gsst OR ssst OR leiden OR secpt OR nnnt OR nnns OR ffsf OR "neurobehavioural scale*" OR pasat OR "serial addition task" OR "still-face" OR "still face" OR "evaluation stress test" OR crest OR "neurobehavioural exam*" OR (social* adj2 (evaluat* OR self OR threat)) OR (public adj2 (speaking OR speech)) OR ((speech OR speaking) adj2 (task OR test))).ab,id,ti,tm,hw.) NOT (abiotic OR acidification OR alfalfa OR algae OR algal OR alligator OR almond OR animal OR apple OR arabidopsis OR arthropod OR avian OR avocado OR bacillus OR bacteria OR bacterial OR bamboo OR barley OR bass OR bean OR bear OR bee OR beet OR beetle OR birch OR bird OR bison OR blueberry OR boar OR bovine OR broccoli OR broiler OR broilers OR buckwheat OR bull OR cabbage OR candida OR carp OR cassava OR cattle OR cavefish OR cereal OR chestnut OR chick OR chicken OR chickpea OR chrysanthemum OR citrus OR coli OR conifer OR coral OR corals OR cotton OR cow OR cowpea OR croaker OR crop OR crops OR crustacean OR cucumber OR dandelion OR dog OR drosophila OR drought OR duckweed OR earthworm OR earthworms OR eel OR elegans OR equine OR eucalyptus OR ewe OR finch OR finches OR fish OR flax OR floral OR flounder OR flowering OR fly OR frog OR fruit OR fungal OR fungi OR fungus OR goat OR grape OR grapevine OR grass OR guppies OR hamster OR herring OR horse OR hyena OR insect OR kelp OR kiwifruit OR leaf OR leaves OR legume OR lettuce OR liverwort OR locust OR longan OR lotus OR macaque OR macaques OR maize OR mangrove OR maples OR marine OR mice OR microalgae OR midge OR millet OR mite OR mollusc OR monkey OR mosquito OR moss OR mouse OR murine OR mycelia OR nutritional OR oak OR osmotic OR ovine OR oxidative OR oyster OR papaya OR passerine OR pea OR peach OR perch OR persimmon OR petunia OR pig OR pigs OR pine OR pistachio OR plant OR pollution OR poplar OR porcine OR poster OR potato OR prawn OR pumpkin OR pylori OR quail OR radish OR rapeseed OR rat OR rats OR reed OR rice OR rodent OR root OR roots OR ruminant OR rye OR ryegrass OR salamander OR salinity OR salmon OR salmonid OR salmonids OR salt OR scallop OR seabass OR seabream OR seagrass OR shark OR sheep OR silkworm OR snail OR soil OR sorghum OR soybean OR sparrow OR sparrows OR sponge OR spruce OR squid OR starling OR stickleback OR strawberry OR sturgeon OR sugarcane OR sunflower OR swine OR switchgrass OR tea OR termite OR tomato OR tree OR trout OR truffle OR urchin OR vegetable OR vegetables OR virus OR vole OR walnut OR watercress OR watermelon OR weed OR whale OR wheat OR whitefly OR yarrow OR yeast OR yellowfin OR zea OR zebrafish OR zinnia).ti,id. NOT (exp animals/ not humans.sh.) AND "Journal Article".dt

***MEDLINE***

((exp "Epigenomics"/ OR exp "DNA Methylation"/ OR (methylation OR dnam OR epigenetic* OR epigenom*).tw,kf.) AND ((cortisol adj2 (reactivity OR response)) OR (("hpa axis" OR hpaa OR "hypothalamic pituitary adrenal") adj5 (function OR response OR reactivity OR regulation)) OR (stress* adj2 (acute OR test OR task OR response OR reactivity OR regulation OR psychological OR psychosocial OR social OR mental OR cortisol)) OR tsst OR trier OR groningen OR gsst OR ssst OR leiden OR secpt OR nnnt OR nnns OR ffsf OR "neurobehavioural scale*" OR pasat OR "serial addition task" OR "still-face" OR "still face" OR "evaluation stress test" OR crest OR "neurobehavioural exam*" OR (social* adj2 (evaluat* OR self OR threat)) OR (public adj2 (speaking OR speech)) OR ((speech OR speaking) adj2 (task OR test))).tw,kf.) NOT (abiotic OR acidification OR alfalfa OR algae OR algal OR alligator OR almond OR animal OR apple OR arabidopsis OR arthropod OR avian OR avocado OR bacillus OR bacteria OR bacterial OR bamboo OR barley OR bass OR bean OR bear OR bee OR beet OR beetle OR birch OR bird OR bison OR blueberry OR boar OR bovine OR broccoli OR broiler OR broilers OR buckwheat OR bull OR cabbage OR candida OR carp OR cassava OR cattle OR cavefish OR cereal OR chestnut OR chick OR chicken OR chickpea OR chrysanthemum OR citrus OR coli OR conifer OR coral OR corals OR cotton OR cow OR cowpea OR croaker OR crop OR crops OR crustacean OR cucumber OR dandelion OR dog OR drosophila OR drought OR duckweed OR earthworm OR earthworms OR eel OR elegans OR equine OR eucalyptus OR ewe OR finch OR finches OR fish OR flax OR floral OR flounder OR flowering OR fly OR frog OR fruit OR fungal OR fungi OR fungus OR goat OR grape OR grapevine OR grass OR guppies OR hamster OR herring OR horse OR hyena OR insect OR kelp OR kiwifruit OR leaf OR leaves OR legume OR lettuce OR liverwort OR locust OR longan OR lotus OR macaque OR macaques OR maize OR mangrove OR maples OR marine OR mice OR microalgae OR midge OR millet OR mite OR mollusc OR monkey OR mosquito OR moss OR mouse OR murine OR mycelia OR nutritional OR oak OR osmotic OR ovine OR oxidative OR oyster OR papaya OR passerine OR pea OR peach OR perch OR persimmon OR petunia OR pig OR pigs OR pine OR pistachio OR plant OR pollution OR poplar OR porcine OR poster OR potato OR prawn OR pumpkin OR pylori OR quail OR radish OR rapeseed OR rat OR rats OR reed OR rice OR rodent OR root OR roots OR ruminant OR rye OR ryegrass OR salamander OR salinity OR salmon OR salmonid OR salmonids OR salt OR scallop OR seabass OR seabream OR seagrass OR shark OR sheep OR silkworm OR snail OR soil OR sorghum OR soybean OR sparrow OR sparrows OR sponge OR spruce OR squid OR starling OR stickleback OR strawberry OR sturgeon OR sugarcane OR sunflower OR swine OR switchgrass OR tea OR termite OR tomato OR tree OR trout OR truffle OR urchin OR vegetable OR vegetables OR virus OR vole OR walnut OR watercress OR watermelon OR weed OR whale OR wheat OR whitefly OR yarrow OR yeast OR yellowfin OR zea OR zebrafish OR zinnia).ti,kf. NOT (exp animals/ not humans.sh.) AND "Journal Article".pt

***Scopus***

(INDEXTERMS(Epigenomics) OR INDEXTERMS("DNA Methylation") OR TITLE-ABS-KEY(methylation OR dnam OR epigenetic* OR epigenom*)) AND TITLE-ABS-KEY((cortisol W/2 (reactivity OR response)) OR (("hpa axis" OR hpaa OR "hypothalamic pituitary adrenal") W/5 (function OR response OR reactivity OR regulation)) OR (stress* W/2 (acute OR test OR task OR response OR reactivity OR regulation OR psychological OR psychosocial OR social OR mental OR cortisol)) OR tsst OR trier OR groningen OR gsst OR ssst OR leiden OR secpt OR nnnt OR nnns OR ffsf OR "neurobehavioural scale*" OR pasat OR "serial addition task" OR "still-face" OR "still face" OR "evaluation stress test" OR crest OR "neurobehavioural exam*" OR (social* W/2 (evaluat* OR self OR threat)) OR (public W/2 (speaking OR speech)) OR ((speech OR speaking) W/2 (task OR test))) AND NOT TITLE(abiotic OR acidification OR alfalfa OR algae OR algal OR alligator OR almond OR animal OR apple OR arabidopsis OR arthropod OR avian OR avocado OR bacillus OR bacteria OR bacterial OR bamboo OR barley OR bass OR bean OR bear OR bee OR beet OR beetle OR birch OR bird OR bison OR blueberry OR boar OR bovine OR broccoli OR broiler OR broilers OR buckwheat OR bull OR cabbage OR candida OR carp OR cassava OR cattle OR cavefish OR cereal OR chestnut OR chick OR chicken OR chickpea OR chrysanthemum OR citrus OR coli OR conifer OR coral OR corals OR cotton OR cow OR cowpea OR croaker OR crop OR crops OR crustacean OR cucumber OR dandelion OR dog OR drosophila OR drought OR duckweed OR earthworm OR earthworms OR eel OR elegans OR equine OR eucalyptus OR ewe OR finch OR finches OR fish OR flax OR floral OR flounder OR flowering OR fly OR frog OR fruit OR fungal OR fungi OR fungus OR goat OR grape OR grapevine OR grass OR guppies OR hamster OR herring OR horse OR hyena OR insect OR kelp OR kiwifruit OR leaf OR leaves OR legume OR lettuce OR liverwort OR locust OR longan OR lotus OR macaque OR macaques OR maize OR mangrove OR maples OR marine OR mice OR microalgae OR midge OR millet OR mite OR mollusc OR monkey OR mosquito OR moss OR mouse OR murine OR mycelia OR nutritional OR oak OR osmotic OR ovine OR oxidative OR oyster OR papaya OR passerine OR pea OR peach OR perch OR persimmon OR petunia OR pig OR pigs OR pine OR pistachio OR plant OR pollution OR poplar OR porcine OR poster OR potato OR prawn OR pumpkin OR pylori OR quail OR radish OR rapeseed OR rat OR rats OR reed OR rice OR rodent OR root OR roots OR ruminant OR rye OR ryegrass OR salamander OR salinity OR salmon OR salmonid OR salmonids OR salt OR scallop OR seabass OR seabream OR seagrass OR shark OR sheep OR silkworm OR snail OR soil OR sorghum OR soybean OR sparrow OR sparrows OR sponge OR spruce OR squid OR starling OR stickleback OR strawberry OR sturgeon OR sugarcane OR sunflower OR swine OR switchgrass OR tea OR termite OR tomato OR tree OR trout OR truffle OR urchin OR vegetable OR vegetables OR virus OR vole OR walnut OR watercress OR watermelon OR weed OR whale OR wheat OR whitefly OR yarrow OR yeast OR yellowfin OR zea OR zebrafish OR zinnia) AND NOT KEY(abiotic OR acidification OR alfalfa OR algae OR algal OR alligator OR almond OR animal OR apple OR arabidopsis OR arthropod OR avian OR avocado OR bacillus OR bacteria OR bacterial OR bamboo OR barley OR bass OR bean OR bear OR bee OR beet OR beetle OR birch OR bird OR bison OR blueberry OR boar OR bovine OR broccoli OR broiler OR broilers OR buckwheat OR bull OR cabbage OR candida OR carp OR cassava OR cattle OR cavefish OR cereal OR chestnut OR chick OR chicken OR chickpea OR chrysanthemum OR citrus OR coli OR conifer OR coral OR corals OR cotton OR cow OR cowpea OR croaker OR crop OR crops OR crustacean OR cucumber OR dandelion OR dog OR drosophila OR drought OR duckweed OR earthworm OR earthworms OR eel OR elegans OR equine OR eucalyptus OR ewe OR finch OR finches OR fish OR flax OR floral OR flounder OR flowering OR fly OR frog OR fruit OR fungal OR fungi OR fungus OR goat OR grape OR grapevine OR grass OR guppies OR hamster OR herring OR horse OR hyena OR insect OR kelp OR kiwifruit OR leaf OR leaves OR legume OR lettuce OR liverwort OR locust OR longan OR lotus OR macaque OR macaques OR maize OR mangrove OR maples OR marine OR mice OR microalgae OR midge OR millet OR mite OR mollusc OR monkey OR mosquito OR moss OR mouse OR murine OR mycelia OR nutritional OR oak OR osmotic OR ovine OR oxidative OR oyster OR papaya OR passerine OR pea OR peach OR perch OR persimmon OR petunia OR pig OR pigs OR pine OR pistachio OR plant OR pollution OR poplar OR porcine OR poster OR potato OR prawn OR pumpkin OR pylori OR quail OR radish OR rapeseed OR rat OR rats OR reed OR rice OR rodent OR root OR roots OR ruminant OR rye OR ryegrass OR salamander OR salinity OR salmon OR salmonid OR salmonids OR salt OR scallop OR seabass OR seabream OR seagrass OR shark OR sheep OR silkworm OR snail OR soil OR sorghum OR soybean OR sparrow OR sparrows OR sponge OR spruce OR squid OR starling OR stickleback OR strawberry OR sturgeon OR sugarcane OR sunflower OR swine OR switchgrass OR tea OR termite OR tomato OR tree OR trout OR truffle OR urchin OR vegetable OR vegetables OR virus OR vole OR walnut OR watercress OR watermelon OR weed OR whale OR wheat OR whitefly OR yarrow OR yeast OR yellowfin OR zea OR zebrafish OR zinnia) AND NOT (INDEXTERMS(animals) AND NOT INDEXTERMS(humans)) AND DOCTYPE(Article)

***Web of Science***

(TS=(methylation OR dnam OR epigenetic* OR epigenom*) AND TS=((cortisol NEAR/2 (reactivity OR response)) OR (("hpa axis" OR hpaa OR "hypothalamic pituitary adrenal") NEAR/5 (function OR response OR reactivity OR regulation)) OR (stress* NEAR/2 (acute OR test OR task OR response OR reactivity OR regulation OR psychological OR psychosocial OR social OR mental OR cortisol)) OR tsst OR trier OR groningen OR gsst OR ssst OR leiden OR secpt OR nnnt OR nnns OR ffsf OR "neurobehavioural scale*" OR pasat OR "serial addition task" OR "still-face" OR "still face" OR "evaluation stress test" OR crest OR "neurobehavioural exam*" OR (social* NEAR/2 (evaluat* OR self OR threat)) OR (public NEAR/2 (speaking OR speech)) OR ((speech OR speaking) NEAR/2 (task OR test)))) NOT (TI=(abiotic OR acidification OR alfalfa OR algae OR algal OR alligator OR almond OR animal OR apple OR arabidopsis OR arthropod OR avian OR avocado OR bacillus OR bacteria OR bacterial OR bamboo OR barley OR bass OR bean OR bear OR bee OR beet OR beetle OR birch OR bird OR bison OR blueberry OR boar OR bovine OR broccoli OR broiler OR broilers OR buckwheat OR bull OR cabbage OR candida OR carp OR cassava OR cattle OR cavefish OR cereal OR chestnut OR chick OR chicken OR chickpea OR chrysanthemum OR citrus OR coli OR conifer OR coral OR corals OR cotton OR cow OR cowpea OR croaker OR crop OR crops OR crustacean OR cucumber OR dandelion OR dog OR drosophila OR drought OR duckweed OR earthworm OR earthworms OR eel OR elegans OR equine OR eucalyptus OR ewe OR finch OR finches OR fish OR flax OR floral OR flounder OR flowering OR fly OR frog OR fruit OR fungal OR fungi OR fungus OR goat OR grape OR grapevine OR grass OR guppies OR hamster OR herring OR horse OR hyena OR insect OR kelp OR kiwifruit OR leaf OR leaves OR legume OR lettuce OR liverwort OR locust OR longan OR lotus OR macaque OR macaques OR maize OR mangrove OR maples OR marine OR mice OR microalgae OR midge OR millet OR mite OR mollusc OR monkey OR mosquito OR moss OR mouse OR murine OR mycelia OR nutritional OR oak

OR osmotic OR ovine OR oxidative OR oyster OR papaya OR passerine OR pea OR peach OR perch OR persimmon OR petunia OR pig OR pigs OR pine OR pistachio OR plant OR pollution OR poplar OR porcine OR poster OR potato OR prawn OR pumpkin OR pylori OR quail OR radish OR rapeseed OR rat OR rats OR reed OR rice OR rodent OR root OR roots OR ruminant OR rye OR ryegrass OR salamander OR salinity OR salmon OR salmonid OR salmonids OR salt OR scallop OR seabass OR seabream OR seagrass OR shark OR sheep OR silkworm OR snail OR soil OR sorghum OR soybean OR sparrow OR sparrows OR sponge OR spruce OR squid OR starling OR stickleback OR strawberry OR sturgeon OR sugarcane OR sunflower OR swine OR switchgrass OR tea OR termite OR tomato OR tree OR trout OR truffle OR urchin OR vegetable OR vegetables OR virus OR vole OR walnut OR watercress OR watermelon OR weed OR whale OR wheat OR whitefly OR yarrow OR yeast OR yellowfin OR zea OR zebrafish OR zinnia)) NOT (AK=(abiotic OR acidification OR alfalfa OR algae OR algal OR alligator OR almond OR animal OR apple OR arabidopsis OR arthropod OR avian OR avocado OR bacillus OR bacteria OR bacterial OR bamboo OR barley OR bass OR bean OR bear OR bee OR beet OR beetle OR birch OR bird OR bison OR blueberry OR boar OR bovine OR broccoli OR broiler OR broilers OR buckwheat OR bull OR cabbage OR candida OR carp OR cassava OR cattle OR cavefish OR cereal OR chestnut OR chick OR chicken OR chickpea OR chrysanthemum OR citrus OR coli OR conifer OR coral OR corals OR cotton OR cow OR cowpea OR croaker OR crop OR crops OR crustacean OR cucumber OR dandelion OR dog OR drosophila OR drought OR duckweed OR earthworm OR earthworms OR eel OR elegans OR equine OR eucalyptus OR ewe OR finch OR finches OR fish OR flax OR floral OR flounder OR flowering OR fly OR frog OR fruit OR fungal OR fungi OR fungus OR goat OR grape OR grapevine OR grass OR guppies OR hamster OR herring OR horse OR hyena OR insect OR kelp OR kiwifruit OR leaf OR leaves OR legume OR lettuce OR liverwort OR locust OR longan OR lotus OR macaque OR macaques OR maize OR mangrove OR maples OR marine OR mice OR microalgae OR midge OR millet OR mite OR mollusc OR monkey OR mosquito OR moss OR mouse OR murine OR mycelia OR nutritional OR oak OR osmotic OR ovine OR oxidative OR oyster OR papaya OR passerine OR pea OR peach OR perch OR persimmon OR petunia OR pig OR pigs OR pine OR pistachio OR plant OR pollution OR poplar OR porcine OR poster OR potato OR prawn OR pumpkin OR pylori OR quail OR radish OR rapeseed OR rat OR rats OR reed OR rice OR rodent OR root OR roots OR ruminant OR rye OR ryegrass OR salamander OR salinity OR salmon OR salmonid OR salmonids OR salt OR scallop OR seabass OR seabream OR seagrass OR shark OR sheep OR silkworm OR snail OR soil OR sorghum OR soybean OR sparrow OR sparrows OR sponge OR spruce OR squid OR starling OR stickleback OR strawberry OR sturgeon OR sugarcane OR sunflower OR swine OR switchgrass OR tea OR termite OR tomato OR tree OR trout OR truffle OR urchin OR vegetable OR vegetables OR virus OR vole OR walnut OR watercress OR watermelon OR weed OR whale OR wheat OR whitefly OR yarrow OR yeast OR yellowfin OR zea OR zebrafish OR zinnia)) NOT (KP=(abiotic OR acidification OR alfalfa OR algae OR algal OR alligator OR almond OR animal OR apple OR arabidopsis OR arthropod OR avian OR avocado OR bacillus OR bacteria OR bacterial OR bamboo OR barley OR bass OR bean OR bear OR bee OR beet OR beetle OR birch OR bird OR bison OR blueberry OR boar OR bovine OR broccoli OR broiler OR broilers OR buckwheat OR bull OR cabbage OR candida OR carp OR cassava OR cattle OR cavefish OR cereal OR chestnut OR chick OR chicken OR chickpea OR chrysanthemum OR citrus OR coli OR conifer OR coral OR corals OR cotton OR cow OR cowpea OR croaker OR crop OR crops OR crustacean OR cucumber OR dandelion OR dog OR drosophila OR drought OR duckweed OR earthworm OR earthworms OR eel OR elegans OR equine OR eucalyptus OR ewe OR finch OR finches OR fish OR flax OR floral OR flounder OR flowering OR fly OR frog OR fruit OR fungal OR fungi OR fungus OR goat OR grape OR grapevine OR grass OR guppies OR hamster OR herring OR horse OR hyena OR insect OR kelp OR kiwifruit OR leaf OR leaves OR legume OR lettuce OR liverwort OR locust OR longan OR lotus OR macaque OR macaques OR maize OR mangrove OR maples OR marine OR mice OR microalgae OR midge OR millet OR mite OR mollusc OR monkey OR mosquito OR moss OR mouse OR murine OR mycelia OR nutritional OR oak OR osmotic OR ovine OR oxidative OR oyster OR papaya OR passerine OR pea OR peach OR perch OR persimmon OR petunia OR pig OR pigs OR pine OR pistachio OR plant OR pollution OR poplar OR porcine OR poster OR potato OR prawn OR pumpkin OR pylori OR quail OR radish OR rapeseed OR rat OR rats OR reed OR rice OR rodent OR root OR roots OR ruminant OR rye OR ryegrass OR salamander OR salinity OR salmon OR salmonid OR salmonids OR salt OR scallop OR seabass OR seabream OR seagrass OR shark OR sheep OR silkworm OR snail OR soil OR sorghum OR soybean OR sparrow OR sparrows OR sponge OR spruce OR squid OR starling OR stickleback OR strawberry OR sturgeon OR sugarcane OR sunflower OR swine OR switchgrass OR tea OR termite OR tomato OR tree OR trout OR truffle OR urchin OR vegetable OR vegetables OR virus OR vole OR walnut OR watercress OR watermelon OR weed OR whale OR wheat OR whitefly OR yarrow OR yeast OR yellowfin OR zea OR zebrafish OR zinnia)) AND (DT=Article)

**Additional Methods for the Meta-Analyses**

The escalc() function in the R package *metafor* was used to estimate Fisher's *r*-to-*z* transformed correlation coefficient with a measure of variance for each effect in the meta-analysis. Each estimate was based on one or more of the following statistics, depending on their availability: Pearson's *r*, *t*, or *p* with *n*. For one study, *t* was obtained by dividing the unstandardised regression coefficient by its standard error [3]. Pearson's *r* was obtained from the estimated Fisher's coefficients using the convert_z2r() function in the R package *esc*. For the two-level meta-analyses (i.e., the basic random effects models), the Hartung-Knapp method was used to compute the confidence intervals, as recommended by IntHout et al. [4]. The benefit of this approach is that it can help to control the rate of false positives, especially when there are few estimates in the meta-analysis [4]. Meta-regression was implemented using the 'mods' argument in the rma.mv() function in *metafor*.

As noted in the main text, a multilevel approach was used to account for the dependency between some of the effects [5]. If multiple effects were reported for different, non-overlapping loci (e.g., amplicons) in the same participants, all effects were included and an additional level was added to account for the dependency. Similarly, if effects for different studies were included from partially overlapping samples, a level was added to the meta-analysis to account for the dependency. If a study reported separate effects for multiple *overlapping* loci for the same set of participants, we included the effect for the largest locus. For example, an effect for the mean level of DNA methylation across a gene took precedence over the effects for the individual CpG sites within the gene. If a study reported effects for a whole cohort with additional subgroup effects, we included the effect from the whole cohort. If effects were only reported for specific subgroups within a cohort (i.e., there was no effect for the full cohort), we used the aggregate.escalc() function in the R package *metafor* to aggregate the subgroup effects into a single estimate before including them in the meta-analysis. If a study reported multiple effects that were comparable apart from their adjustment for different variables, the least-adjusted effect was included in the primary meta-analysis and – where possible – sensitivity analyses were performed with the most-adjusted effects (i.e., those that adjusted for the highest number of additional variables).

As noted in the main text, we computed additional estimates where possible using the publicly available data set GSE77445 [2]. The estimates were based on pre-processed Illumina Infinium HumanMethylation 450K BeadChip DNA methylation data and a pre-computed TSST-G cortisol AUCi variable [2]. Following a recommendation by Du et al. [6], the beta values in the public data set were converted to M values (with an offset of 0.001) prior to analysis, as they better approximate a normal distribution. Following a recommendation by van Rooij et al. [7], linear regression was used to obtain the estimates (rather than limma or some other alternative). The dependent variable was cortisol AUCi, predicted from the mean level of DNA methylation across all of the probes that were annotated to the gene plus age and sex, which were included as covariates. Additional effects were computed for *NR3C1*, *SLC6A4*, and *FKBP5*, referred to as “Balfour et al. (2025)” in the relevant forest plots.

### Additional Meta-Analyses

#### Figure S1

Association Between *NR3C1* Methylation and the Cortisol Response (Recovery)

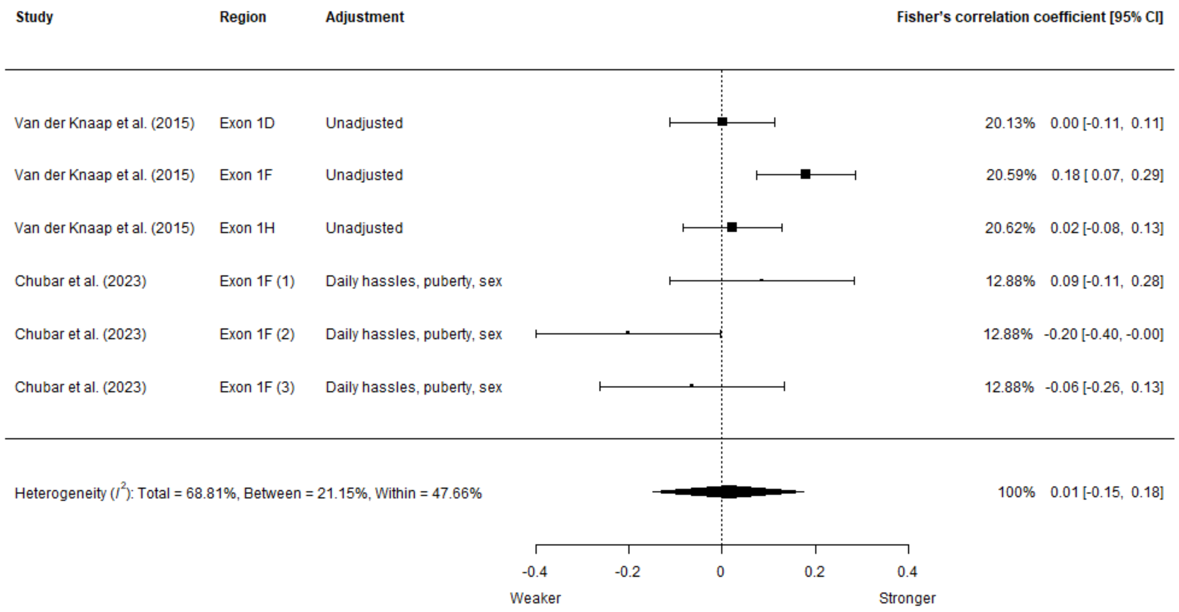


*Note*. Pooled estimate: *r* = .01 (95% CI = -.15, .17), *p* = .84. Weaker = weaker (slower) cortisol recovery, Stronger = stronger (faster) cortisol recovery. Region column indicates the specific area within *NR3C1*, if the authors reported on more than one for the same participant group. Adjustment column indicates which variables were adjusted for. Additional level included in the meta-analysis to account for the lack of independence between estimates for different genomic loci within studies.

**Figure S2**

Association Between *NR3C1* Methylation and the Cortisol Response (AUCg)

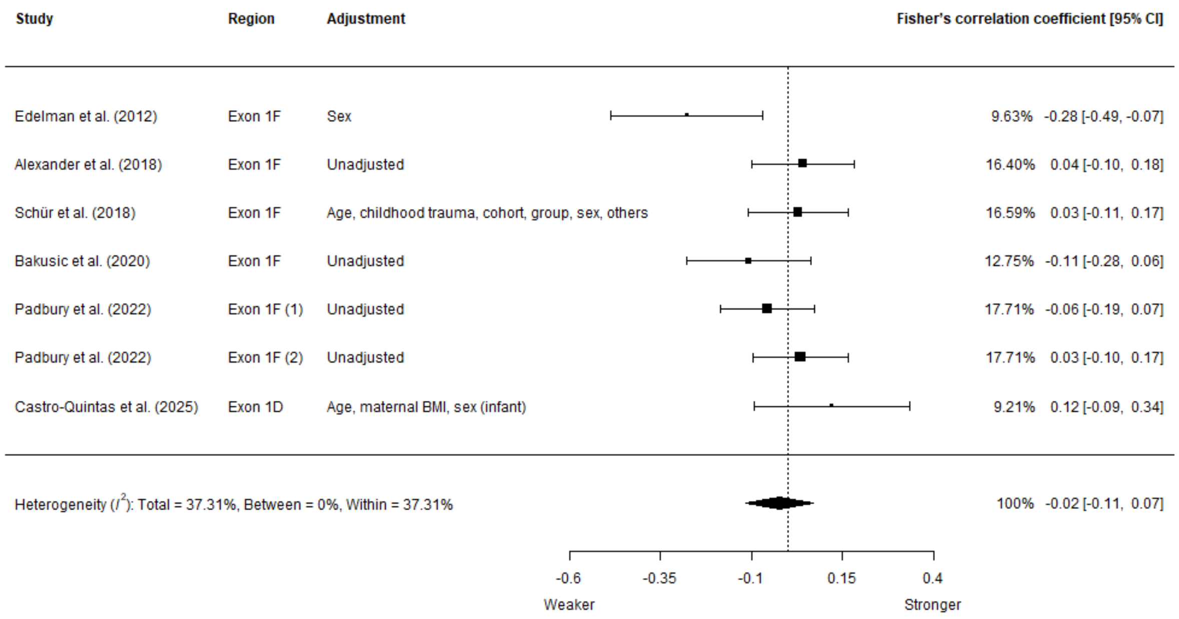


*Note*. Pooled estimate: *r* = -.02 (95% CI = -.11, .07), *p* = .59. Region column indicates the specific area within *NR3C1*, if the authors reported on more than one for the same participant group. Adjustment column indicates which variables were adjusted for, with the word "infant" in brackets to indicate an infant-only sample. Additional level included in the meta-analysis to account for the lack of independence between estimates for different genomic loci within studies.

**Figure S3**

Association Between *NR3C1* Methylation and the Cortisol Response (AUCi)

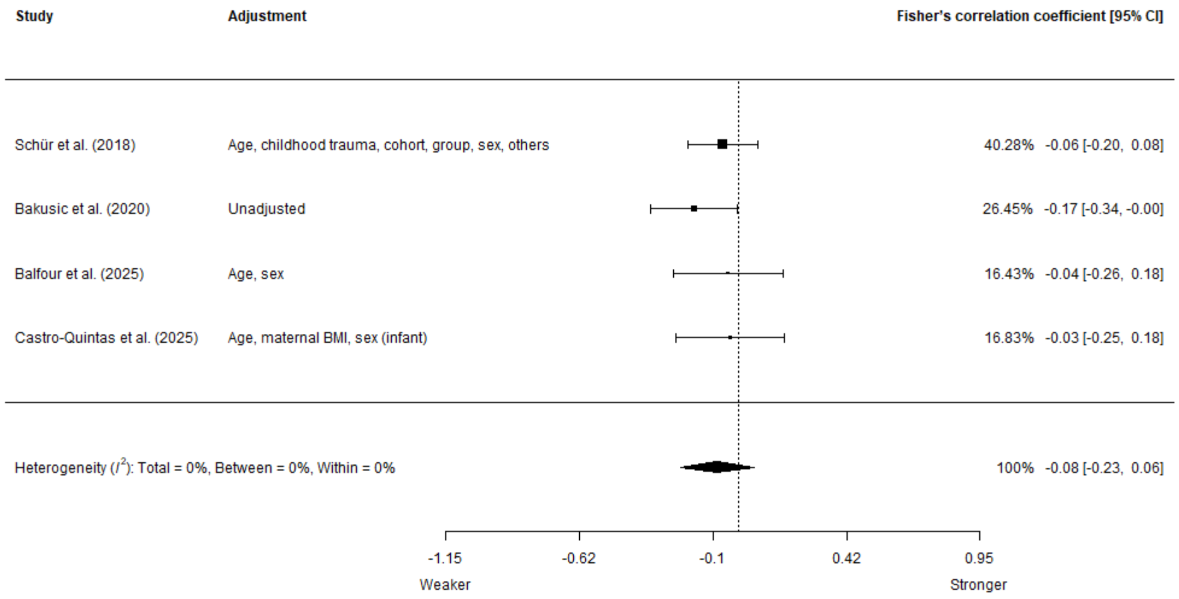


*Note*. Pooled estimate: *r* = -.08 (95% CI = -.22, .06), *p* = .16. Adjustment column indicates which variables were adjusted for, with the word "infant" in brackets to indicate an infant-only sample. Additional level included in the meta-analysis to account for partially overlapping cohorts between studies.

**Figure S4**

Association Between *SLC6A4* Methylation and the Cortisol Response (AUCg)

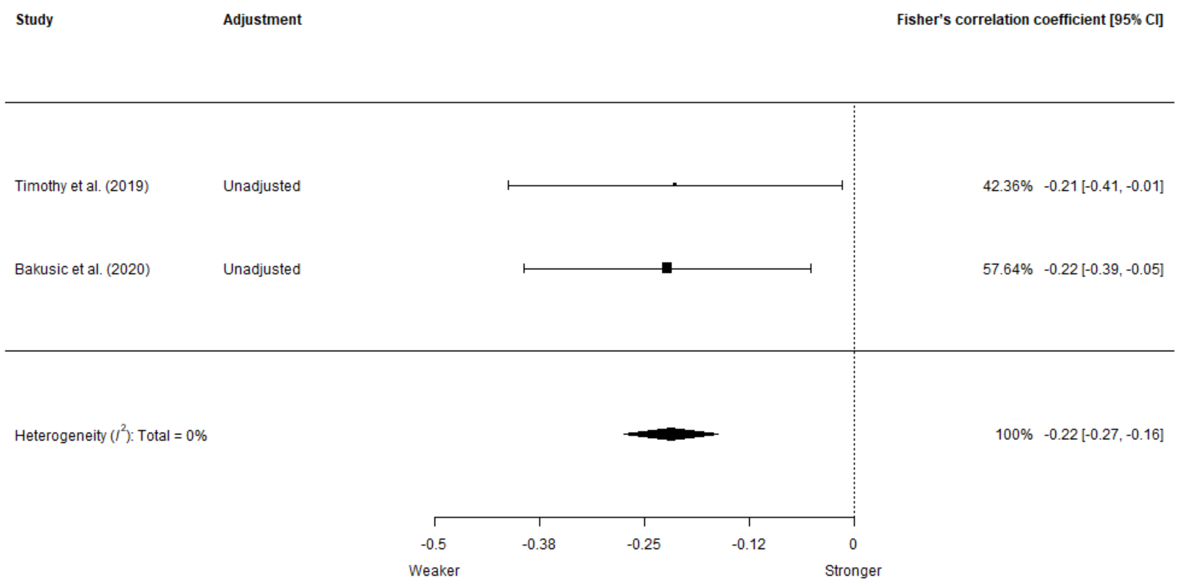


*Note*. Pooled estimate: $r$ = -.21 (95% CI = -.27, -.16), $p$ = .01. Adjustment column indicates which variables were adjusted for.

**Figure S5**

Association Between *FKBP5* Methylation and the Cortisol Response (AUCi)

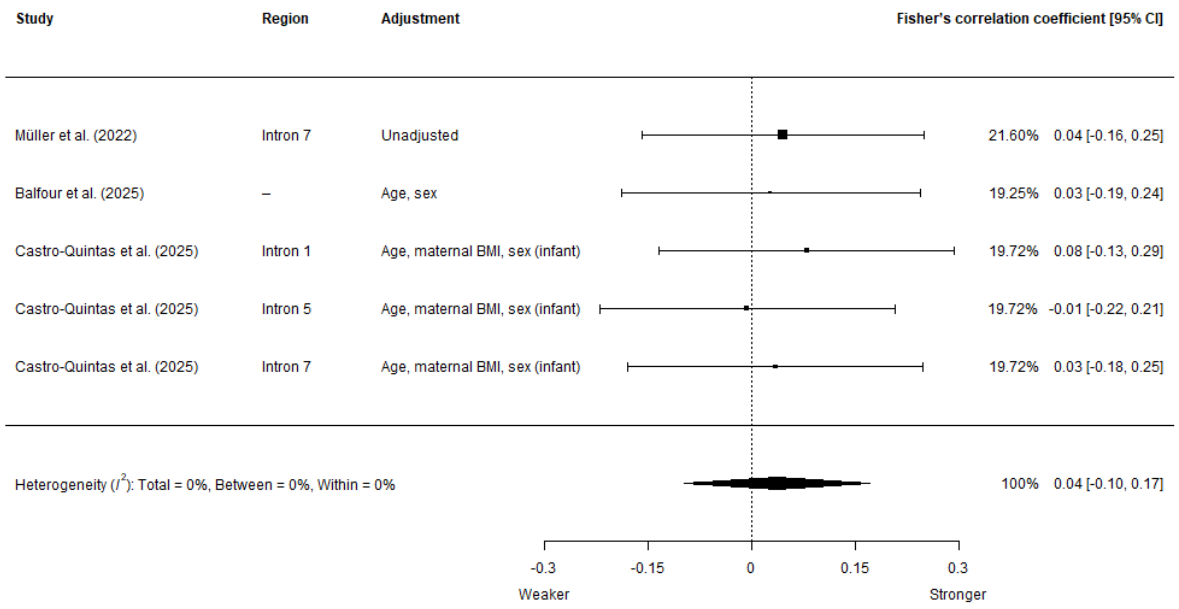


*Note*. Pooled estimate: *r* = .04 (95% CI = -.1, .17), *p* = .5. Weaker = weaker cortisol response, Stronger = stronger cortisol response. Region column indicates the specific area within *FKBP5*, if the authors reported on more than one for the same participant group. Adjustment column indicates which variables were adjusted for, with the word “infant” in brackets to indicate an infant-only sample. Additional level included in the meta-analysis to account for the lack of independence between estimates for different genomic loci within studies.

**Figure S6**

Association Between *FKBP5* Methylation and the Cortisol Response (AUCg)

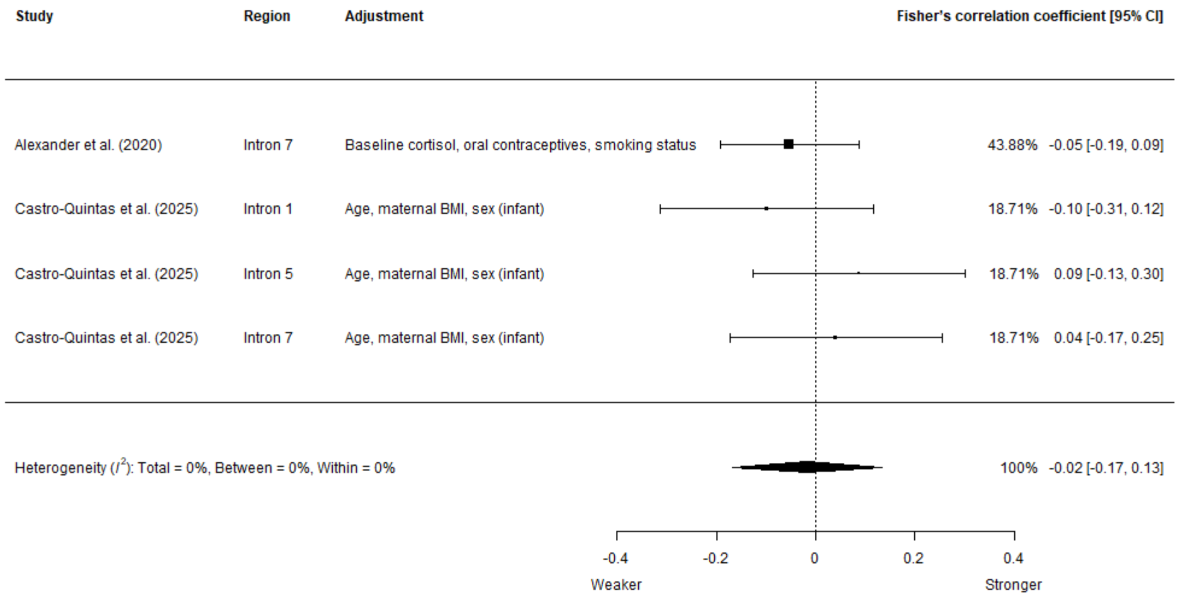


*Note*. Pooled estimate: *r* = -.02 (95% CI = -.17, .13), *p* =.73. Weaker = weaker cortisol response, Stronger = stronger cortisol response. Region column indicates the specific area within *FKBP5*, if the authors reported on more than one for the same participant group. Adjustment column indicates which variables were adjusted for, with the word “infant” in brackets to indicate an infant-only sample. Additional level included in the meta-analysis to account for the lack of independence between estimates for different genomic loci within studies.

**Figure S7**

Association Between *KITLG* Methylation (cg27512205) and the Cortisol Response (AUCi)

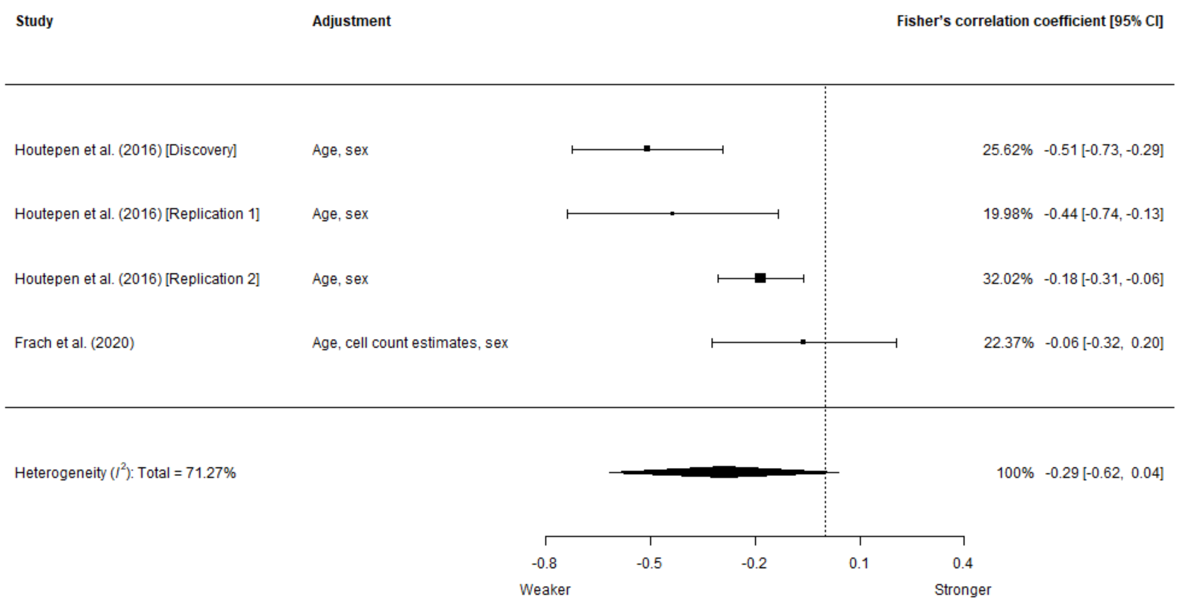


*Note*. Pooled estimate: $r$ = -.28 (95% CI = -.55, .04), $p$ =.07. Weaker = weaker cortisol response, Stronger = stronger cortisol response. Adjustment column indicates which variables were adjusted for.

**Figure S8**

Indirect Effect of Childhood Trauma on the Cortisol Response (AUCi) Through *KITLG* Methylation (cg27512205)

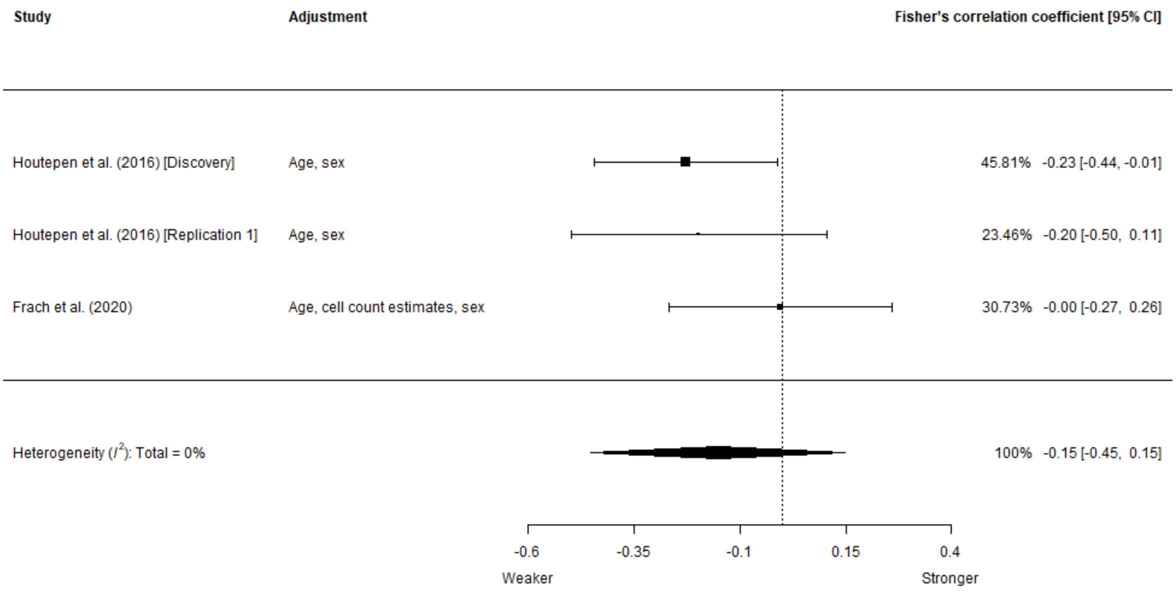


*Note*. Pooled estimate: *r* = -.15 (95% CI = -.42, .15), *p* = .16. Adjustment column indicates which variables were adjusted for.

**Figure S10**

Adjusted Association Between *NR3C1* Methylation and the Cortisol Response (Recovery)

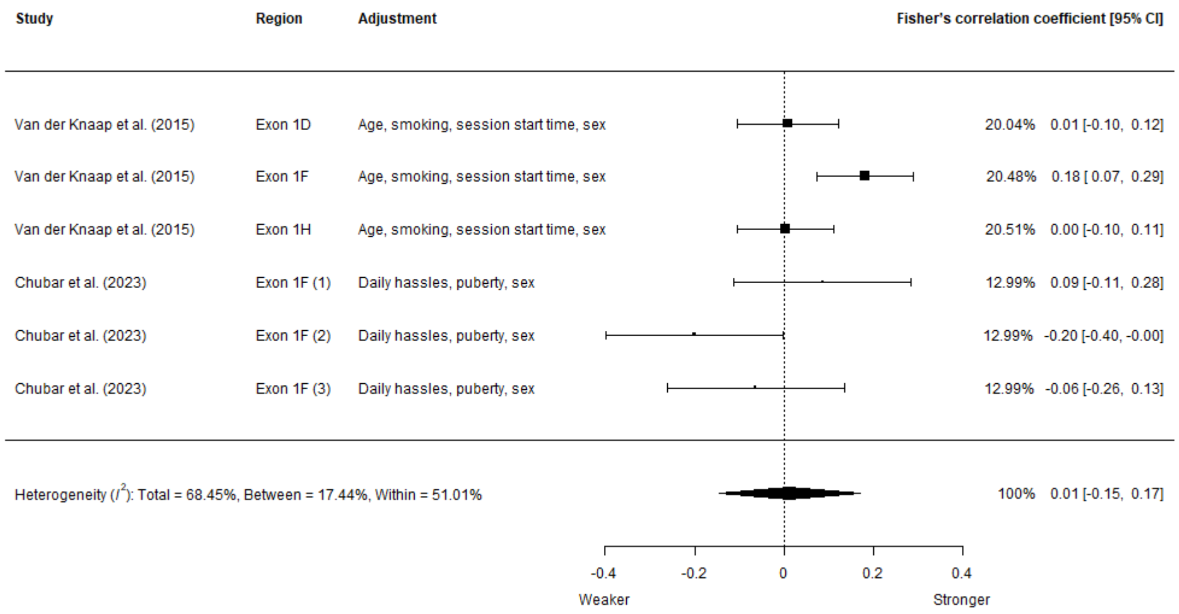


*Note*. Pooled estimate: $r$ =.01 (95% CI = -.14,.17), $p$ =.85. Weaker = weaker (slower) cortisol recovery, Stronger = stronger (faster) cortisol recovery. Region column indicates the specific area within *NR3C1*, if the authors reported on more than one for the same participant group. Adjustment column indicates which variables were adjusted for. Additional level included to account for the lack of independence between estimates for different genomic loci within studies. Least-adjusted effects have been replaced with their most-adjusted equivalents where possible.

**Figure S11**

Adjusted Association Between *KITLG* Methylation (cg27512205) and the Cortisol Response (AUCi)

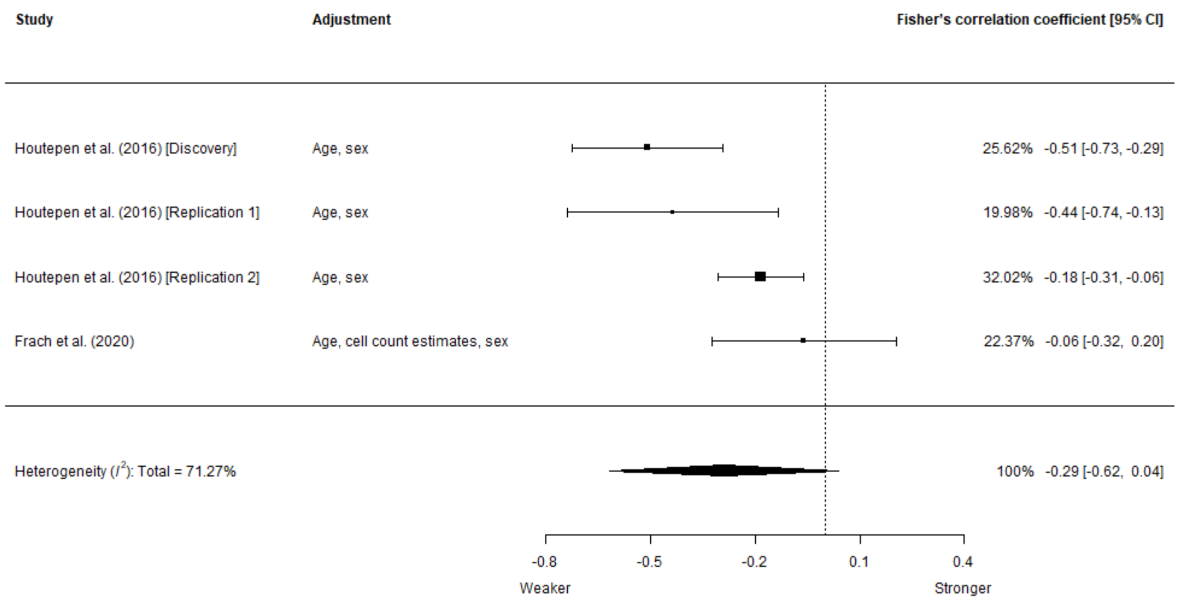


*Note*. Pooled estimate: $r$ = -.28 (95% CI = -.55, .04), $p$ =.07. Weaker = weaker cortisol response, Stronger = stronger cortisol response. Adjustment column indicates which variables were adjusted for. Least-adjusted effects have been replaced with their most-adjusted equivalents where possible.

**Figure S12**

Adjusted Association Between *NR3C1* Methylation and the Cortisol Response (Reactivity)

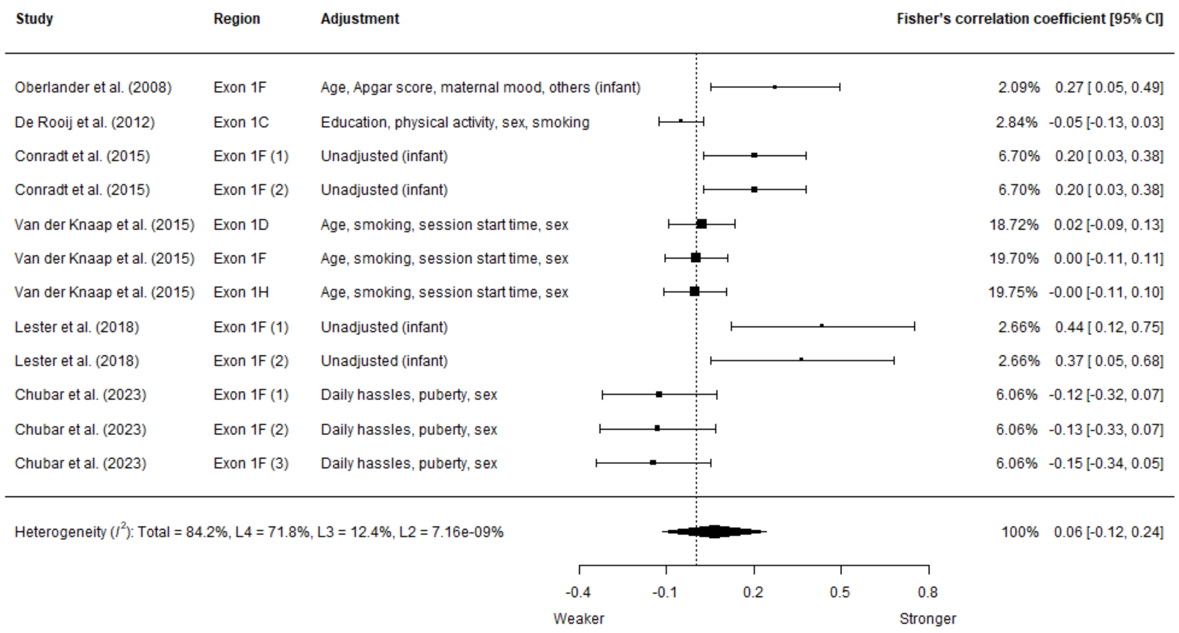


*Note*. Pooled estimate: $r$ =.06 (95% CI = -.12, .24), $p$ =.45. Weaker = weaker cortisol response, Stronger = stronger cortisol response. Region column indicates the specific area within *NR3C1*, if the authors reported on more than one for the same participant group. Adjustment column indicates which variables were adjusted for, with the word “infant” in brackets to indicate an infant-only sample. Additional levels included in the meta-analysis to account for the lack of independence between estimates for different genomic loci within studies and partially overlapping cohorts between studies. Least-adjusted effects have been replaced with their most-adjusted equivalents where possible.

**Figure S13**

Association Between *NR3C1* Exon $1_F$ Methylation and the Cortisol Response (Reactivity)

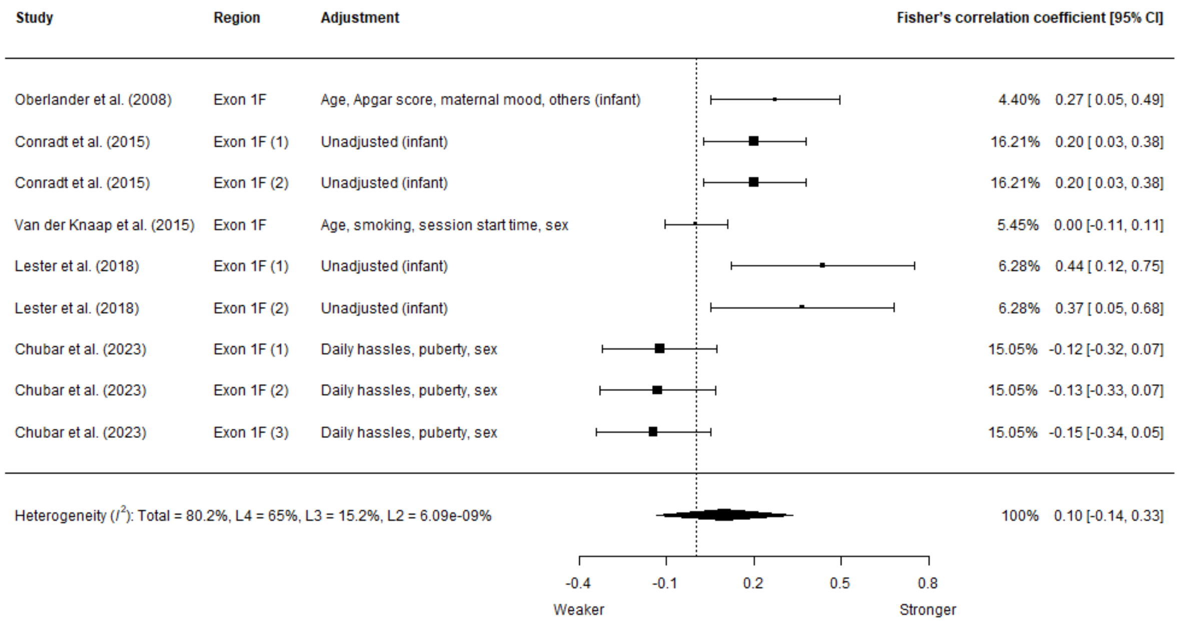


*Note*. Pooled estimate: $r$ = .09 (95% CI = -.16, .32), $p$ = .44. Weaker = weaker cortisol response, Stronger = stronger cortisol response. Region column indicates the specific area within *NR3C1*, if the authors reported on more than one for the same participant group. Adjustment column indicates which variables were adjusted for, with the word "infant" in brackets to indicate an infant-only sample. Additional levels included in the meta-analysis to account for the lack of independence between estimates for different genomic loci within studies and partially overlapping cohorts between studies.

**Figure S14**

Association Between *NR3C1* Exon $1_F$ Methylation and the Cortisol Response (Recovery)

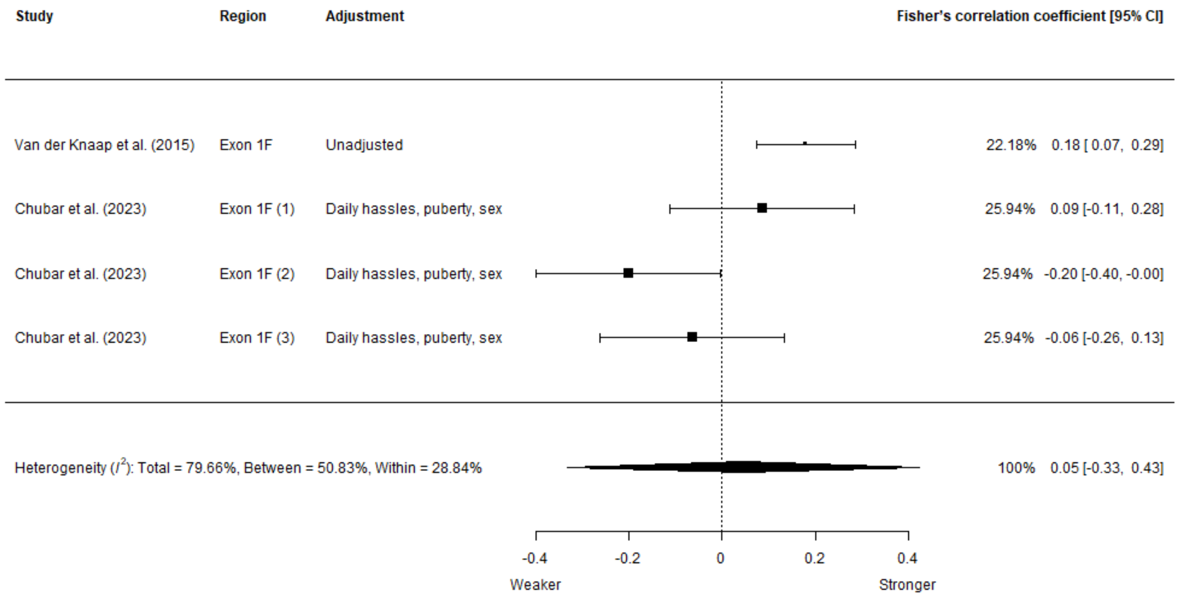


*Note*. Pooled estimate: *r* =.05 (95% CI = -.32, .4), *p* =.72. Weaker = weaker (slower) cortisol recovery, Stronger = stronger (faster) cortisol recovery. Region column indicates the specific area within *NR3C1*, if the authors reported on more than one for the same participant group. Adjustment column indicates which variables were adjusted for. Additional levels included in the meta-analysis to account for the lack of independence between estimates for different genomic loci within studies.

**Figure S15**

Association Between *NR3C1* Exon $1_F$ DNA Methylation and the Cortisol Response (AUCg)

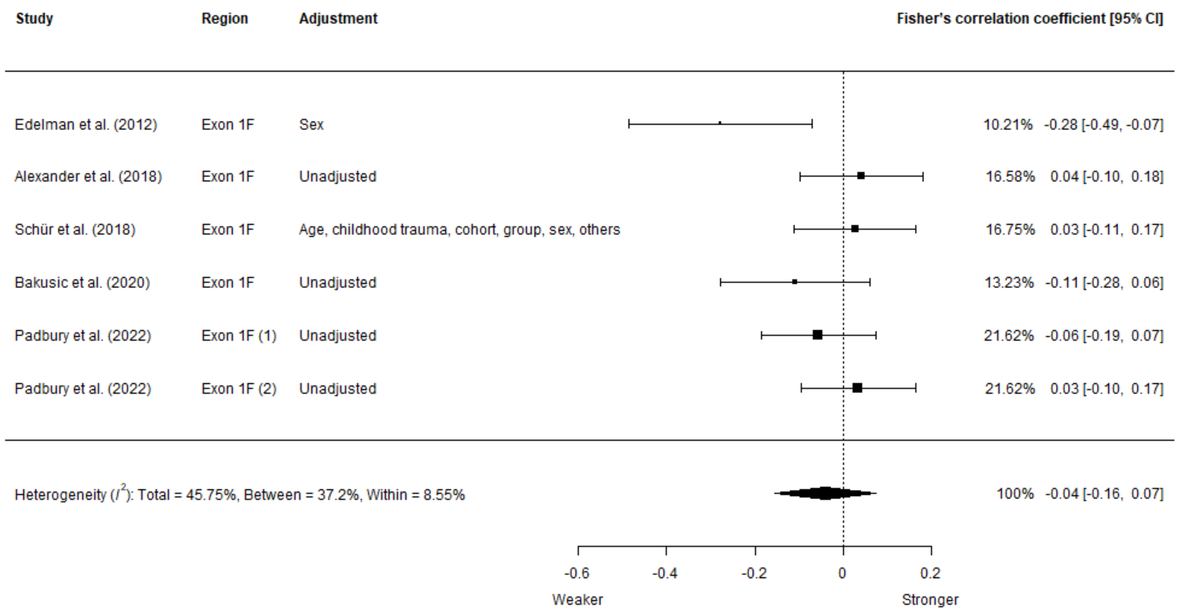


*Note*. Pooled estimate: *r* = -.04 (95% CI = -.15, .07), *p* = .4. Weaker = weaker cortisol response, Stronger = stronger cortisol response. Region column indicates the specific area within *NR3C1*, if the authors reported on more than one for the same participant group. Adjustment column indicates which variables were adjusted for. Additional level included in the meta-analysis to account for the lack of independence between estimates for different genomic loci within studies.

**Figure S16**

Risk of Bias Judgements

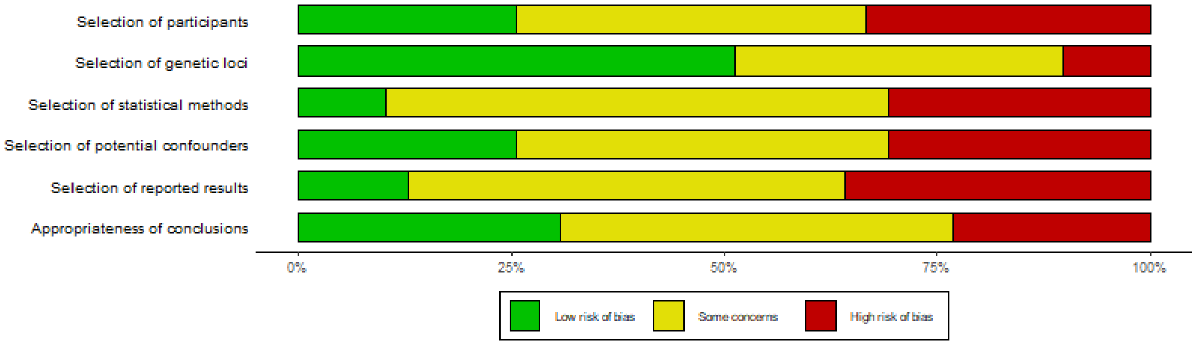


*Note*. Bars represent the percentage of studies assigned a given risk of bias judgement for each domain.

**Methylome-Wide Power Analysis**

We used the R package *EpipwR* [8] to perform methylome-wide power analyses. Power was defined as the proportion of true differentially methylated positions that were detected after adjustment for the false discovery rate. We assumed a true population effect size equal to the mean absolute estimate ($r$ = .2497) for the 15 032 CpG sites that were nominally associated (at α = .05) with AUCi in the discovery data investigated by Houtepen et al. (2016) (GSE77445; we conducted the analysis using linear regression and M values, with no covariates). The standard deviation was set to the standard deviation for the estimates (*SD* = 0.0325). Power analyses were performed for five different sample sizes, based on the two largest independent methylome-wide studies (*N* = 85 [9] and *N* = 58 [10]), the largest study based on methylome-wide data (*N* = 298) [11], and the largest study in the review more generally (*N* = 675) [12]. Power analyses assumed different tissues types and different counts of true differentially methylated positions (i.e., CpG sites with a true non-zero correlation with AUCi). At 85 participants, estimated power ranged from just .0025 (assuming 100 differentially methylated positions) to .0205 (assuming 6 400 differentially methylated positions) in whole blood (Table S3).

**Additional Comments on Heterogeneity**

***Analysis of the Cortisol Response***

The most common outcomes were AUCi (*k* = 19), reactivity (*k* = 14) and AUCg (*k* = 9). Also, three studies included a measure representing the recovery of the HPA axis after exposure to the stressor. Only four studies adjusted for baseline cortisol as a covariate.

***Tissue Type***

The studies investigated DNA methylation in different tissue types. The most common tissue was whole blood (*k* = 19), followed by buccal cells (*k* = 9), placenta (*k* = 6), and saliva (*k* = 4). Each tissue has a different function and pattern of DNA methylation, so it could be that some studies were unable to replicate previous findings because they investigated a different tissue. One study could not replicate a previously-reported effect for *KITLG* [10], but they used $CD14^+$ monocytes rather than whole blood or buccal cells, which were used in the original study [9]. Another study [13] was unable to replicate a previously reported association between *SLC6A4* methylation and the cortisol response that varied by *5-HTTLPR* genotype [14], although they used peripheral blood mononuclear cells instead of whole blood [13]. Similarly, one study [15] reported no effect of *FKBP5* methylation by genotype subgroup in whole blood, although an earlier study had reported evidence for an interaction between the two using cord blood leukocytes [11]. Also consistent with an effect of tissue type, one study reported evidence for an effect of *NR3C2* methylation in placenta but not in buccal cells for the same participants [16]. It may be that the association of interest cannot be observed reliably in peripheral tissue. Unfortunately, it may not be feasible to study more theoretically relevant tissues such as the hippocampus (which facilitates negative feedback within the HPA axis) in the present context.

***Stress Protocols***

The studies used different stress protocols, with most of the non-infant studies (*k* = 29) implementing a version of the Trier Social Stress Test (TSST). In its standard form, the TSST includes a speech task followed by a mental arithmetic task. Both tasks are performed in front of a panel, to create social-evaluative threat [17]. Three of the five non-infant studies that used an alternative protocol also included a speech task. Among the infant studies (*k* = 10), three used a version of the Face-to-Face Still-Face (FFSF) paradigm, which involves the caregiver suddenly stopping interaction with the infant and maintaining a neutral facial expression. Three studies used the Neonatal Intensive Care Unit Network Neurobehavioral Scales (NNNS), which includes interaction with a stranger and exposure to visual and auditory stimuli. Two studies used Ainsworth's Strange Situation procedure (SSP), which includes a phase where the caregiver leaves the infant. The different protocols may have contributed to the mixed results, as they may vary in their capacity to elicit a cortisol response [17].

***Control for Potential Confounders***

The studies adjusted for different variables. Nine studies did not adjust for any potential confounders. To investigate the possible effect of confounding, we performed sensitivity analyses that replaced the least-adjusted effects with their most-adjusted equivalents where possible. The pooled estimates for *NR3C1* and recovery (Figure S10) and *KITLG* and AUCi (Figure S11) did not change substantially after adjustment. The effect for *NR3C1* and reactivity decreased, consistent with some potential confounding (*r* changed from .09 to .06 [95% CI = -.12, .24], *p* from .27 to .45; Figure S12, *k* = 12, *N* = 1 389). The additional variables that were adjusted for were education, smoking, physical activity, experimental session start time, and sex.

***Timing of the DNA Sample***

The timing of the DNA sample varied between the studies. Five studies clearly indicated that the sample was obtained within 24 hours of the stressor [18, 19, 3, 20, 21] and seven studies reported that it was obtained between 1 month and 2 years before the stressor [22, 23, 16, 11, 24, 25, 26]. For the most part, the other studies were not as clear as they could have been about the duration of time between the sample and the stressor. It is possible that the duration between the two affects the strength or nature of the relationship, so it should be reported on clearly in any future research. Studies were also often unclear about whether the sample was obtained before or after the stressor. This could be an important detail because patterns of DNA methylation are most likely affected by the stressor itself [27, 20], potentially limiting direct comparison between studies that obtained the DNA sample before and after the stressor. We encourage researchers to clearly report on the timing of the DNA sample in relation to the stressor.

***Methods Used to Compute the DNA Methylation Variables***

The studies varied in the DNA methylation variables they used. Several studies analysed the mean level of DNA methylation across selected CpG sites or units in a candidate region, with or without secondary analyses for the individual sites or units. One study used the sum instead of the mean [3], one study analysed a unique predictor based on the number of CpG sites with more than 0% DNA methylation [21], two studies applied the median split to a continuous DNA methylation variable [14, 28], and two studies examined co-methylated factors [13, 29]. The various methods used to generate DNA methylation variables for analysis may have contributed to the mixed results, and they are a potentially somewhat arbitrary statistical decision that should ideally be pre-specified in a published research protocol.

**Other Comments**

***Researcher Degrees of Freedom***

Decisions made during data analysis and at other points in a research project are sometimes referred to as researcher degrees of freedom [30]. These decisions can lead to a hidden multiple testing problem, where researchers can perform a wide range of analyses and only report on or focus on those that are consistent with their hypothesis, inflating the rate of type one error [31]. In some cases, it may be that the effect of interest can only be observed in a specific analytical situation. For example, when adjusting for a particular set of covariates, exploring a particular interaction or subgroup effect, using a particular threshold to dichotomise a variable, using a particular method to analyse the cortisol response (e.g., AUCg rather than AUCi), or using a specific composite DNA methylation variable (e.g., a particular co-methylated factor or the mean level of DNA methylation for a specific set of CpG sites). The effect may then fail to replicate in subsequent studies because it reflects noise in the original data rather than a genuine relationship in the population, which may account for some of the mixed findings in the reviewed literature. To mitigate possible problems with researcher degrees of freedom, we encourage researchers to report extensive sensitivity analyses that demonstrate the robustness of their results to key analytical decisions, such as the choice of covariates, the exclusion of participants, and the method that was used to obtain a predictor variable from the DNA methylation data.

***Protocol Recommendations***

We encourage researchers to publish a protocol on OSF before they run their analyses. Ideally, the protocol should include the following information, at minimum: which criteria will be used to determine the loci that will be taken forward for analysis, which variables will be controlled for and how (e.g., age will be controlled for through residualisation and sex will be included as a covariate), which DNA methylation variables will be computed and how (e.g., the mean level of DNA methylation across a particular set of CpG sites), how cortisol will be analysed (e.g., AUCg and AUCi, adjustment for baseline cortisol), and a description of any planned moderation, mediation, or subgroup analyses. Together, sensitivity analyses and pre-published protocols may help to mitigate any potential issue with researcher degrees of freedom.
